# Integrated genomics identify novel immunotherapy targets for malignant mesothelioma

**DOI:** 10.1101/2020.01.23.20018523

**Authors:** Anca Nastase, Amit Mandal, Shir Kiong Lu, Hima Anbunathan, Deborah Morris-Rosendahl, Yu Zhi Zhang, Xiao-Ming Sun, Spyridon Gennatas, Robert C Rintoul, Matthew Edwards, Alex Bowman, Tatyana Chernova, Tim Benepal, Eric Lim, Anthony Newman Taylor, Andrew G Nicholson, Sanjay Popat, Anne E Willis, Marion MacFarlane, Mark Lathrop, Anne M Bowcock, Miriam F Moffatt, William OCM Cookson

## Abstract

**Background:** Malignant pleural mesothelioma (MPM) is an aggressive malignancy with limited effective therapies.

**Methods:** In order to identify therapeutic targets, we integrated SNP genotyping, sequencing and transcriptomics from tumours and low-passage patient-derived cells.

**Results:** Previously unrecognised losses of *SUFU* locus (10q24.32), observed in 21% of 118 tumours, resulted in disordered expression of transcripts from Hedgehog pathways and the T-cell synapse including *VISTA*. Co-deletion of Interferon Type I genes and *CDKN2A* was present in half of tumours and was a predictor of poor survival. We also found previously unrecognised deletions in *RB1* in 26% of cases and show sub-micromolar responses to downstream PLK1, CHEK1 and Aurora Kinase inhibitors in primary MPM cells. Defects in Hippo pathways that included *RASSF7* amplification and *NF2* or *LATS1*/2 mutations were present in 50% of tumours and were accompanied by micromolar responses to the YAP1 inhibitor Verteporfin.

**Conclusions:** Our results suggest new therapeutic avenues in MPM and provide targets and biomarkers for immunotherapy.

## Introduction

Malignant pleural mesothelioma (MPM) is an aggressive malignancy associated with asbestos exposure. Global mesothelioma deaths are estimated to be 38,400 each year^1^. MPM shows limited responses to all treatments. Although 20% of tumours may transiently regress after checkpoint immunotherapy^2,3,^ PD-L1 is expressed at a low level in the majority of MPM cases^3^ and predictors of response are unknown. The molecular landscape is not complex but to date known recurrent lesions have not yet defined effective therapeutic targets^4,5^.

Intense fibrosis invariably accompanies MPM, causing intractable pain and dyspnoea. For example, in the UK MesobanK tumour repository^6^ 65% of MPM have <25% of tumour cells visible on surgical biopsy and only 8% of MPM comprise >75% malignant cells. It is likely therefore that tumour-matrix interactions are cardinal features of the disease.

Inflammation and fibrosis in the pleura are normally adaptive mechanisms that seal off foci of injury or infection. Incomplete macrophage phagocytosis of inhaled high-aspect ratio asbestos fibres induces sustained inflammation and cytokine release^7,8^, increased mesothelial proliferation, oxidative DNA damage^9^ and double strand DNA breakages^10^ that can cause malignant transformation.

Previous genomic analyses of MPM have shown a mutational landscape dominated by loss of function mutations in *BAP1* and *NF2*^4,5^. Larger structural variations in MPM are common^11^, and recurrent deletions are recognised for *CDKN2A* (located at chromosome 9p21.3), *NF2* (22q12) and *BAP1* (3p21.3).

Given the proclivity for asbestos to induce DNA damage, we extended genomic findings in 121 MPM tumours by fine mapping of copy-number alterations (CNAs) with high-density SNP arrays. We explored the mutational spectrum with whole exome sequencing (WES) in 50 subjects (21 of which had paired blood samples for germline DNA), before extending mutation detection to all tumours with a 57-gene targeted capture next-generation sequencing (TC-NGS) panel (Supplementary File 1_Figure 1a, b and Supplementary File 1_Table 1). In addition, we whole-genome sequenced (WGS) 19 low-passage primary mesothelioma derived cell cultures (PMCC)^12^.

## Results

### Demographic and clinical characteristics

One hundred and five of the 121 patients (87%) were male (Supplementary File 1_Table 2). Ninety tumours exhibited the epithelioid subtype of MPM, 25 were biphasic and 6 were sarcomatoid (Supplementary File 1_Figure1c). Patients with sarcomatoid disease were older than the other two groups (*P*= 0.05). Asbestos exposure had been documented clinically in 69% of cases. The median overall survival (OS) for all subjects was 9.9 months with sarcomatoid patients showing a worse outcome than others, (*P*=0.065) (survival time from diagnosis to death or last follow-up was available for 110 patients; Supplementary File 1_Table 2 and Supplementary File 1_Figure 1d; Supplementary File 2_Table 1), as described^13^.

Given the wide variability between MPM malignant cells and matrix fibrosis, we estimated the tumour content of all samples with ASCAT analysis of the >950K SNPs in the Illumina Infinium OmniExpressExome-8 v1.3 and v1.4 panels. We found the median tumour content was 0.44 (range 0.2-1.0; 1^st^ quartile 0.32; 3^rd^ quartile 0.9; median absolute deviation 0.22).

### Copy number alterations (CNAs) analysis shows recurrent *CDKN2A, RB1* and *SUFU* deletion and *RASSF7* amplification

We analysed the SNPs for CNAs using the GISTIC program^14^. The program estimates genomic boundaries for recurrent CNA events and assigns statistical significance after false discovery rate (FDR) corrections.

Deletion of *CDKN2A* was the most frequent event observed in our samples, detected in 71/118 tumours (60%), with 58 deletions (82%) predicted to be homozygous (Figures 1a and 2). As previously reported^15^, *CDKN2A* loss was associated with worse OS compared with *CDKN2A* wild type patients (8.8 vs 13.0 months, Kaplan-Meier *P*=0.02) (Figure 1c), and with increased copy number burden (Supplementary File 2_Table 5A).

**Figure 1.**
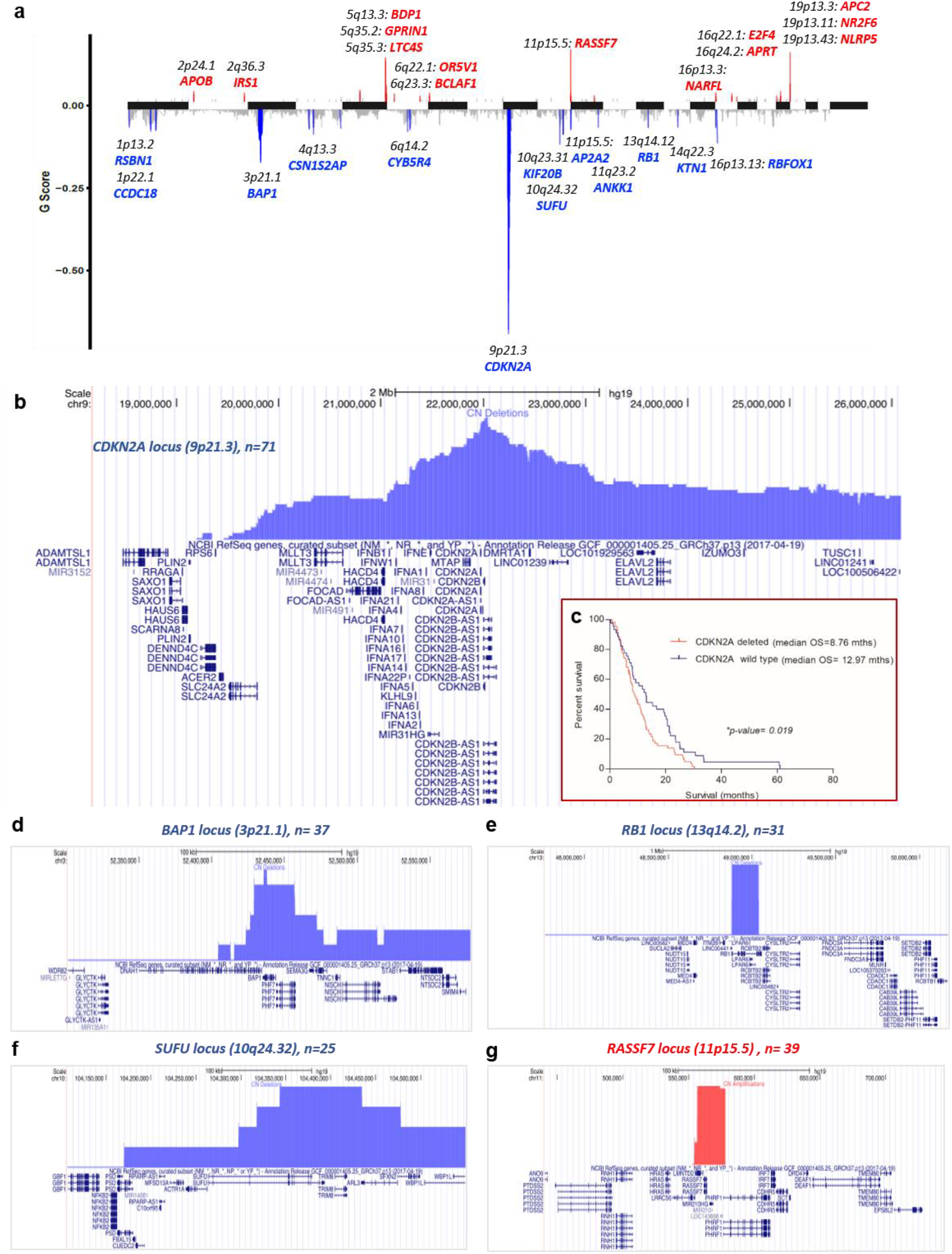
Mapping of copy number alterations in subjects with Malignant Pleural Mesothelioma. a) Statistically significant regions of amplification and deletion from GISTIC analysis of 118 subjects. Peak regions which pass both the G-Score (derived from amplitude and frequency) and q-bound (<0.05) threshold cut-offs are shown for deletions (blue) and amplifications (red) (see also Supplementary File 1_Table 3 and Supplementary File 2_Table 5B for the extensive list of copy-number coordinates for each sample); b) Detailed map of the *CDKN2A* locus using the UCSC Genome Browser (hg19), showing histogram representation of overlap among deletion segments from the 118 subjects. *IFN* Type I genes are commonly within the deleted segments; c) Kaplan-Meier survival analysis of patients with and without *CDKN2A* locus deletions; d-f) Similar Genome Browser based maps of the *BAP1, RB1* and *SUFU* deleted segments; g) Map for amplification segments from the *RASSF7* locus.

**Figure 2.**
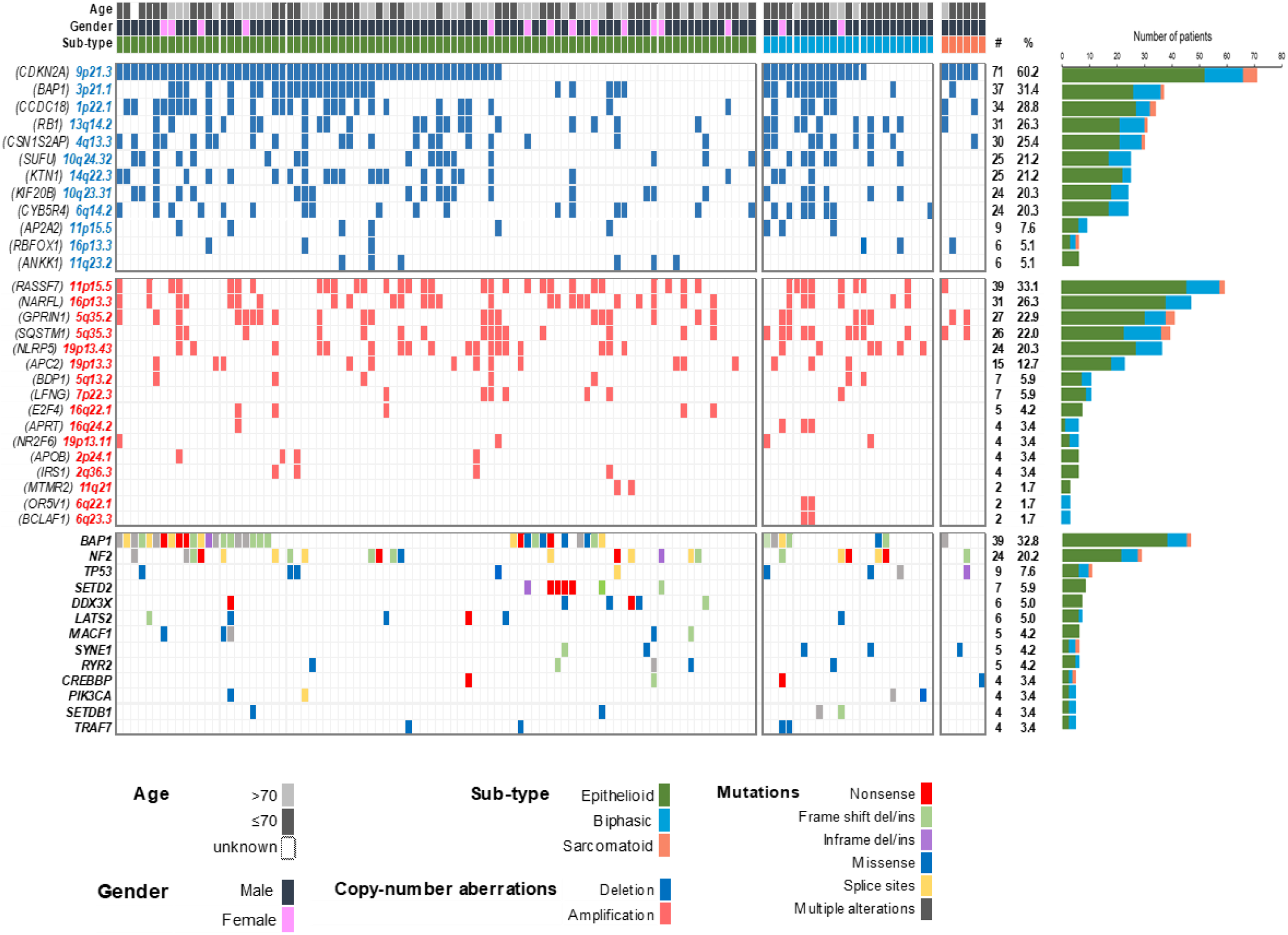
Common genetic alterations in Malignant Pleural Mesothelioma. The most common deletions (top panel), amplifications (middle panel) and mutations (bottom panel) are shown in 118 subjects. CNA analyses are derived from SNP arrays and mutated genes from the targeted capture sequencing panel.

Deletion of the *CDKN2A* region is extensive in many malignancies and a full locus map (Figure 1b) revealed large deletions to also be present in MPM. The map revealed a frequent loss of the closely neighbouring Type I Interferon (IFN) genes, as we have first reported^16^ and has later been confirmed^17^ (Figure 1b and Supplementary File 2_Table 6): 38/118 patients (32%) had predicted homozygous *IFN* Type I loss and 24/118 (20%) had heterozygous loss. The median survival of patients with co-deletion of *CDKN2A* and *IFN* Type I genes was not statistically different to *CDKN2A* deletions alone (8.3 months compared to 10.7, *P*=0.6; Supplementary File 2_Table 6).

We observed frequent deletions at multiple other loci (Figure 1d-f, Supplementary File 1_Table 3 and Supplementary File 2_Table 5B for the extensive list of copy-number coordinates for each sample). The most common novel deletion was the *RB1* locus on 13q14.2 in 31/118 patients (26%). The *RB1* tumour suppressor is activated downstream of CDK4 and CDK6^18^ and its loss predicts a mechanism for resistance to CDK4/6 inhibitors.

Loss of a locus on 10q24.32 containing *SUFU* (*Suppressor of Fused), ARL3 (ADP Ribosylation Factor Like GTPase 3)* and *TRIM8 (Tripartite Motif Containing 8)* was observed in 25/118 tumours (21%). *SUFU* inhibits activation of GLI transcription factors in the Hedgehog pathway, which is known to be disordered during MPM carcinogenesis^19,20^. *TRIM8* acts as a tumour suppressor inducing cell cycle arrest in a TP53 dependant manner, and as an oncogene activating NF-kβ and TNF-α ^21^. It is involved both in immunity and cancer^22^.

Previously unrecognised regions of amplification (Figures 1a and 2, Supplementary File 1_Table 3 and Supplementary File 2_Table 5B for the extensive list of copy-number coordinates for each sample) included a locus on 11p15.5, amplified in 39/118 tumours (33%) that contained *RASSF7* and *miR-210* (Figure 1g). When up-regulated *RASSF7* controls cell growth and apoptosis in different tumours^23^, and functions as an oncogene in NSCLC, interacting with *MST1* to dysregulate Hippo signalling^24^.

Other substantial amplifications included 19q13.43 in 24/118 tumours (20%), containing *NLRP5, ZNF444* and *ZNF787*; 5q35.2 in 27/118 tumours (23%), containing *GPRIN1* immediately adjacent to *CDHR2* which may moderate contact inhibition of epithelial cell growth^25^; and 5q35.3 in 26/118 tumours (22%) containing *LTC4S* and *SQSTM1*. The latter encodes p62, a mediator of autophagy influencing tumorigenesis, malignant growth and resistance to therapy^26^.

We sought replication of the novel CNAs in 98 tumours from the landmark study of Bueno *et al*.^5^ which carried out WES and concurrent RNA-sequencing. We confirmed amplification of *RASSF7* and deletion of *RB1* and *SUFU*, each of which correlated with its transcript abundance (Supplementary File 1_Figure 4 and Supplementary File 2_Table 7).

### Loss of *SUFU* locus, component of Hedgehog signalling pathway, is associated with downregulation of immune related genes

Loss of the *SUFU* locus in 25/118 tumours (21%) was associated with marked upregulation of the *Patched 2* tumour suppressor (*PTCH2*) (Table 1). *Ptch2* overexpression has been observed in *Sufu* knockout mice and is indicative of aberrant Hedgehog signalling^20^. Hedgehog pathways are activated in MPM patients, in the absence of obvious mutations^19^. Also, upregulated were *NHS, HOXA7* and *TRPS1*, each of which regulate tissue differentiation (Table 1).

**Table 1.**
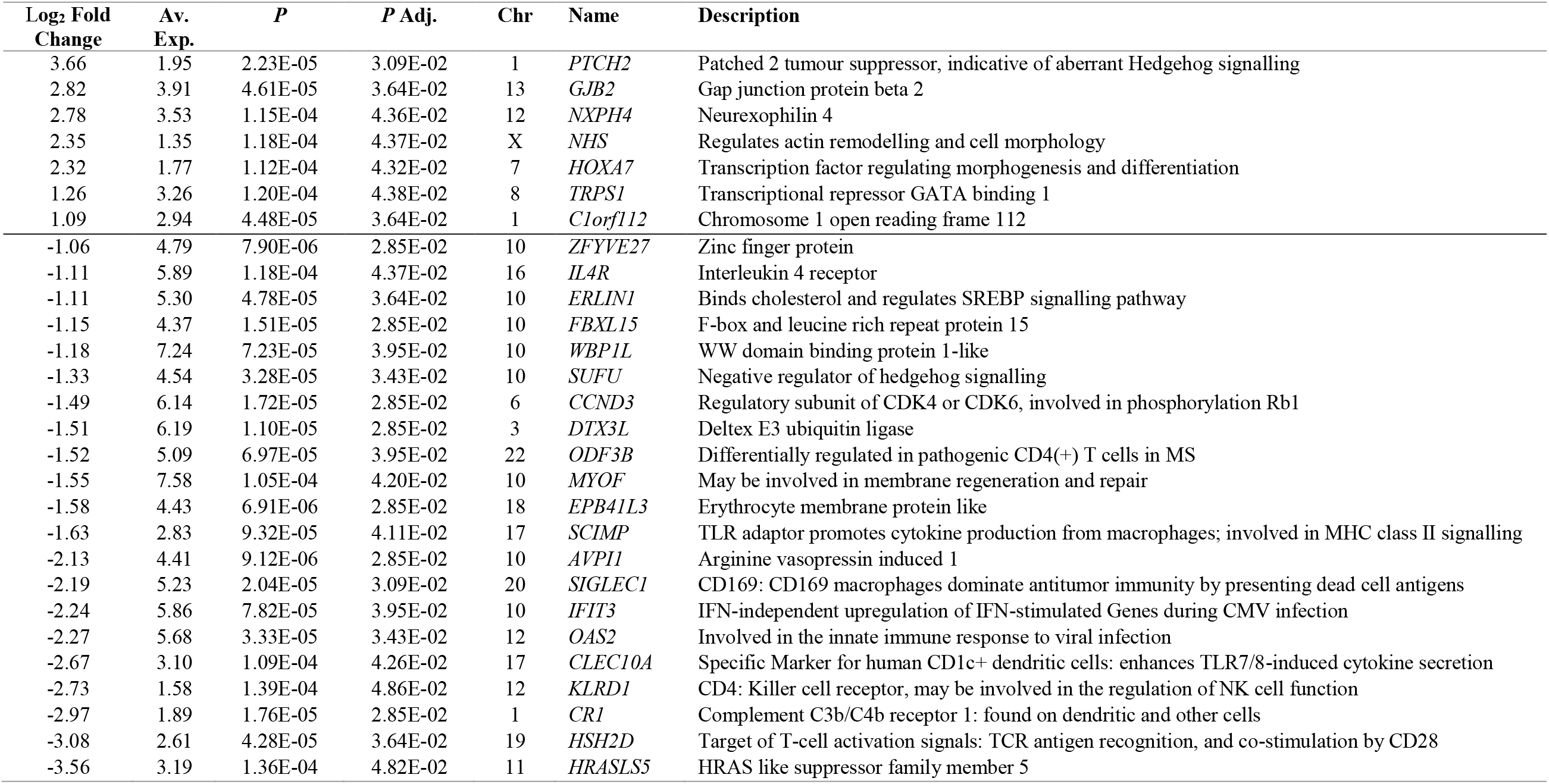
Genes differentially regulated with *SUFU* locus deletions

We found a SMO (Smoothed) inhibitor (Vismodegib, GDC-0499) to be inhibitory in only one PMCC (Figure 4h). Vismodegib has previously been shown to be effective in modulating MPM tumour and stromal interactions in a rat model of MPM^27^, suggesting beneficial effects of Hedgehog inhibition would be worthy of investigation in mixed cellular cultures of human MPM.

*SUFU* loss unexpectedly correlated with downregulation of prominent T-cell genes (Table 1), including *ODF3B*; the killer-cell receptor *KLRD1* (*CD94*); and *HSH2D*, a target of T-cell activation. Downregulated monocyte/macrophage and dendritic cell markers included *IL4R, SCIMP, SIGLEC1* (*CD169*), *CLEC10A*, and *CR1* (Complement C3b/C4b receptor 1).

We confirmed these results in the large independent dataset from Bueno *et al*.^5^, confirming that abundances of Hedgehog pathway transcripts *SUFU, PTCH1* and *PTCH2* correlated with *KLRD1* and *CR1* (Supplementary File 1_Table 5).

### Hippo pathway is deregulated in 50% of MPM patients due to *RASSF7* amplification and *NF2, LATS1/2* mutations

Hippo monitors external factors that shape tissue structure^28^. *NF2* recruits core Hippo signalling pathway members (*LATS1/2*) to inhibit activation of the transcriptional cofactors YAP1 and TAZ^29^. *RASSF7* also regulates Hippo pathways, and its overexpression promotes phosphorylation and nuclear translocation of YAP1^24^. We found *RASSF7* amplification in 39 MPMs, *NF2* mutations in 24, *LATS2* mutations in 6 and *LATS1* in 2, so that non-overlapping lesions in Hippo pathways were present in 52/121 MPMs (43%) and a further 9 MPM had more than one lesion (total 50%) (Figure 2). *WNT5B* transcription, which we found increased in sarcomatoid tumours, may also induce YAP/TAZ activation through non-canonical pathways^30^.

We did not find significant differences in transcript abundances when comparing RNA-sequencing derived transcriptomes for *RASSF7* amplifications to other tumours; or for lesions in Hippo signalling genes (*NF2, LATS1, LATS2)* singly or combined; or for MPMs with or without *SETD* mutations. We did not detect *RASSF7* amplification in any of 19 primary cell lines examined by WGS and SNP array (Supplementary File 1_Figure 5), which may reflect selection in culture for MPM genotypes that grow independently of a fibroblast matrix.

### Investigation of somatic mutations confirms the major driver genes in MPM

WES in 50 tumours (mean coverage 136X; 21 with paired peripheral blood lymphocyte DNA (PBL)) revealed no major loci beyond those previously described^4,5^ (Supplementary File 2_Table 2). We successfully completed TCS-NGS in 119 patients (Supplementary File 2_Table 3 and Supplementary File 1_Figure 1b), 77 of which had paired PBL, achieving a mean coverage of 792X for tumours and 802X for PBL. *BAP1* was mutated in 39 subjects (33%); *NF2* in 24 subjects (20%); *TP53* in 9 subjects (8%) and *SETD2* in 7 subjects (6%) (Figure 2 and Supplementary File 1_Figure 2). Mutations were scattered across coding regions of these genes (Supplementary File 1_Figure 3 b-d), consistent with their putative role as tumour suppressors.

We found two *NRAS* mutations at known oncogenic RAS hotspots (G12V and Q61H). Both mutations were found in sarcomatoid subtype tumours that did not have alterations in *CDKN2A, BAP1* or *NF2*. In TCGA cohort, only one patient with biphasic histology had a G12C *KRAS* hotspot mutation with a concurrent *TP53* mutation but without alterations in other mesothelioma drivers, similar with our cohort. In Bueno *et al*.^5^ cohort also only one patient with sarcomatoid subtype had a Q61K *NRAS* mutation but this was detected simultaneous with a *BAP1* frameshift mutation. We also identified three other RAS pathway related genes by WES: a *NF1* stop mutation (c.6439C>T, p.Q2147*), a splice site *RASA1* mutation (c.829-1_858.del) and a *HRAS* in-frame deletion (c.187_189del, p.E63del).

*TP53* mutations carried a worse prognosis compared with *TP53* wild-type counterparts (mean OS 5.7 vs. 13.6 months, *P*=0.0005), as previously described^5^. We did not detect significant associations of other mutations with survival.

Combined analysis of CNA, WES and TCS-NGS (Figure 2) showed *CDKN2A* deletion to be present in 60% of tumours; *BAP1* mutated or deleted in 54%; *RASSF7* amplification in 33%; *RB1* deleted or mutated in 26%; *NF2* mutated in 20%; *TP53* mutated in 8%; *SETD2* in 6%; *DDX3X* in 5% and *LATS2* in 5%.

We detected a missense germline mutation localized in the UCH domain of *BAP1* from one patient with epithelioid subtype (Supplementary File 1_Figure 3b). In other subjects, single deleterious germline mutations were found in *MSH5* and *MSH6* (representing the mismatch-repair (MMR) pathway), *RB1, SETD6* and *BRCA2*.

### *BAP1* is the most common mutated gene in MPM and is associated with up-regulation of *RET*

We explored the effects of genetic alteration by comparing RNA-sequencing samples with and without specific genetic alterations. When compared to other tumours, *BAP1* mutations or deletions were associated (*P*_*adjusted*_<0.05) with up-regulation of the *RET* proto-oncogene ^31^ and *NNAT*, is the latter associated with poor outcome in multiple cancers ^32^ (Supplementary File 1_Table 6). We replicated the negative association of *BAP1* with *RET* to be present also in the Bueno *et al*.^5^ (r=-0.32, *P*=2.2E-06) and TCGA-Meso^4^ (r=-0.45, *P*=1.3E-06) datasets.

### Mutational signatures 1 and 3 are prevalent in MPM

A median of 31 non-synonymous somatic mutations per tumour exome were present in the 21 WES paired samples, consistent with the low rate observed by Bueno *et al*.^5^. We observed a similar low tumour mutational burden in the 77 paired samples that underwent targeted capture sequencing (Supplementary File 1_Figure 3i).

One patient (NCMR035) had a hypermutated tumour (167 somatic mutations from WES) (Supplementary File 1_Figure 6), accompanied by a frame-shift deletion in *MSH6* (p.Phe1104LeufsX11) and a frame-shift insertion in *PALB2* (p.Met1049AspfsX4). *PALB2* encodes a protein that recruits BRCA2 and RAD51 at the site of double-strand breaks^33^ and plays a critical role in homologous recombination repair.

The mutation spectrum was characterized by C>T transitions, in both WES and TCS-NGS panel data (Supplementary File 1_Figure 3e and 3a respectively), consistent with earlier reports^4,5^. Analysis of mutational signatures ^34,35^ found COSMIC signatures 1 and 3 to be prevalent in the 21 paired WES samples (Supplementary File 1_Figure 3e, f) and in WGS from 19 PMCC^12^ (Supplementary File 1_Figure 3g, h).

Signature 3 is indicative of DNA damage and failed breakpoint repair^35^. In other cancers, signature 3 mutations often accompany biallelic inactivation of *BRCA1* or *BRCA2*, where the inability to repair DNA predicts good responses to platinum therapy. MPM responds poorly to such therapies, and we hypothesise that signatures of DNA damage may follow the actions of asbestos in the progenitor neoplastic cell.

### *WNT5B* is higher expressed in sarcomatoid tumours

We did not see any significant associations between common lesions and histological subtypes. RNA-sequencing however revealed differential transcription between histologies (Supplementary File 1_Table 4). As reported previously^5^, *WNT5B* had higher expression in sarcomatoid tumours. Other genes significantly upregulated (*P*_*adjusted*_<0.001) in non-epithelioid tumours included *GPR176* which acts as a circadian pacesetter^36^, and known adverse factors for other cancers such as *IGF2BP1, CCBE1, HS3ST3A1, TRAM2* and *SERTAD2*.

### High level of VISTA is frequent in epithelioid MPM and its expression level correlates with Hedgehog and immune pathway components

We tested how the most frequent genomic alterations, *BAP1* mutation and *CDKN2A* deletion, were translated at protein levels by staining a subset of 28 tumours (Figure 3a) with antibodies against BAP1 and MTAP (as a potential surrogate marker for *CDKN2A* deletion) (Figure 3d, e).

**Figure 3.**
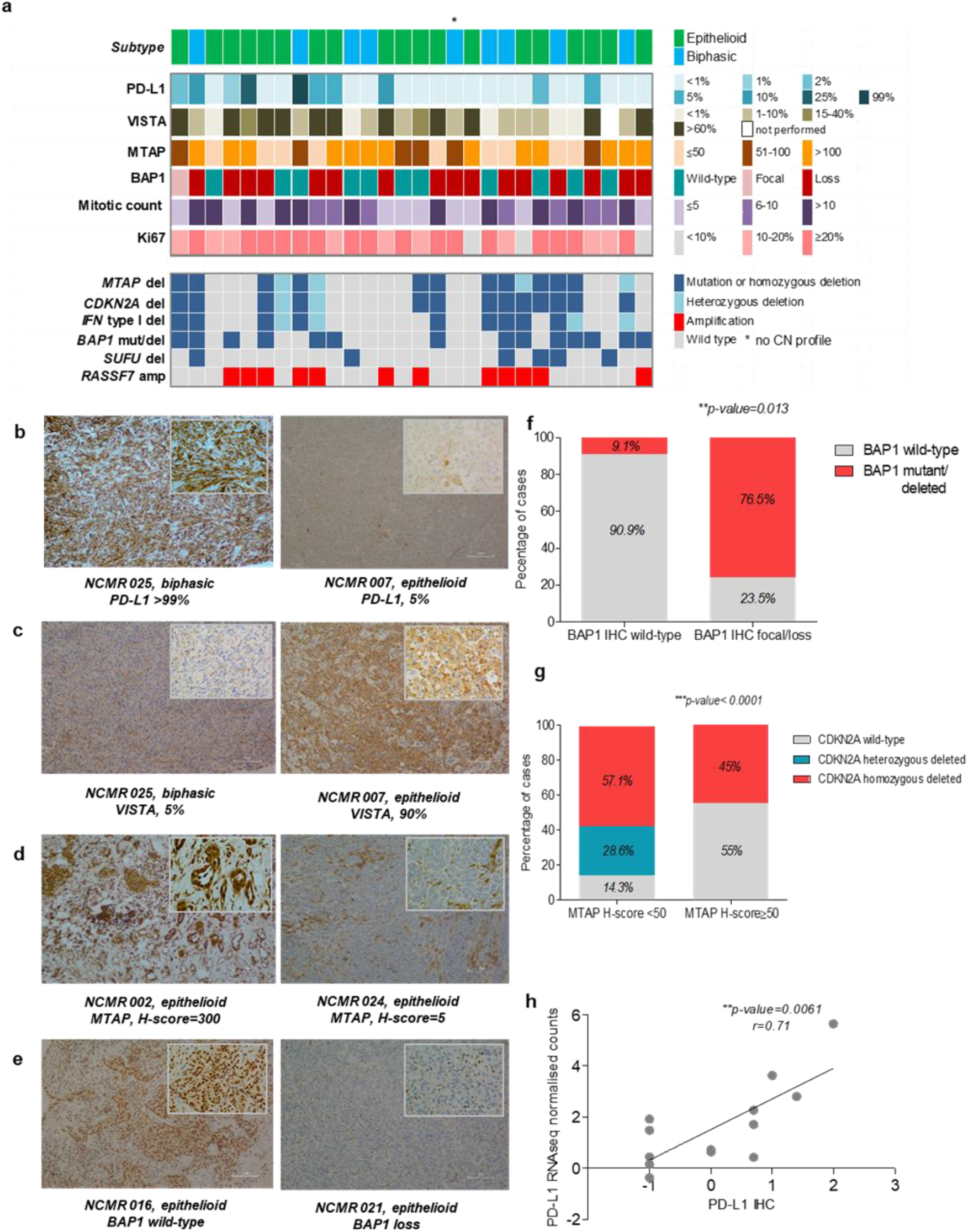
Histologic features of MPM. a) Oncoplot of IHC staining and common genomic alterations; b-e) representative IHC images for PD-L1, MTAP and BAP1 (100X) ; f) analysis on BAP1 genomic and proteomic status by Fishers’ exact test; g) analysis of MTAP H-score in *CDKN2A/MTAP* deleted vs *CDKN2A/MTAP* wild-type by Fisher’s exact t-test, with a threshold for MTAP H-score set at 50; h) Spearman correlation between PD-L1 gene expression and HC.

BAP1 staining revealed general or focal loss in 17/28 (61%) of cases, which only partially associated with *BAP1* mutation or deletion (*P*=0.01) (Figure 3f), as previously suggested^37^. There was significant difference when comparing MTAP H-score between *CDKN2A*/*MTAP* deleted and wild-type samples (*P*=0.001) (Figure 3g). The mitotic count and Ki-67 (both indicators of proliferation) correlated with each other (*P*<0.0001, r=0.42). Ki67 correlated with copy number burden (*P*=0.03, r=0.42) and with MTAP score (*P*=0.04, r=0.39), consistent with disordered cellular division accompanying *CDKN2A* loss.

Checkpoint inhibitors targeting PD-1 and its ligand PD-L1 cause marked tumour regression in some patients with MPM^3^. However, PD-L1 is expressed at a low level, if at all, in most MPM cases and its status imperfectly predicts response to immune checkpoint inhibitors^3^.

In our tumours, IHC staining for PD-L1 was also low, with only 4/28 cases (14%) exhibiting ≥10% expression, including one case >70%. There was a good correlation between PD-L1 (SP263) staining and transcript abundance (*P*<0.01, r=0.7) (Figure 3h). We did not see a consistent relationship between any Hedgehog-related transcripts and PD-L1 in transcriptomic data (Supplementary File 1_Table 5).

High-level staining of the alternative immune-checkpoint protein VISTA (V-domain Ig suppressor of T cell activation)^38^ has been observed in epithelioid MPM, and implies a better prognosis^4,39^. We confirmed a high level of VISTA by IHC in our samples (Figure 3a), and in RNA-sequencing data replicable associations were seen between VISTA and *SUFU, PTCH1, PTCH2, KLRD1* and *CR1* (Supplementary File 1_Table 5).

### Drug-testing shows that primary cell models of MPM are sensitive to cell cycle targeted drugs

We explored potential therapeutic pathways that had been suggested by our genomic findings by determining the half maximal inhibitory concentration (IC_50_) of selected compounds with three PMCC^12^ that had been whole-genome sequenced (Supplementary File 1_ Figure 5). We assessed by Western blots if deletions or mutations of the main MPM drivers were translated to protein levels (Figure 4a). For comparison, we included an immortalized mesothelioma cell line (H2052), a lung adenocarcinoma cell line (A549), and a transformed normal mesothelial cell line (Met-5A). We exposed cells to a range of drug concentrations (0.0005 to 50 micromolar (µM)) using as controls DMSO treated cells (Figure 4b-g).

**Figure 4.**
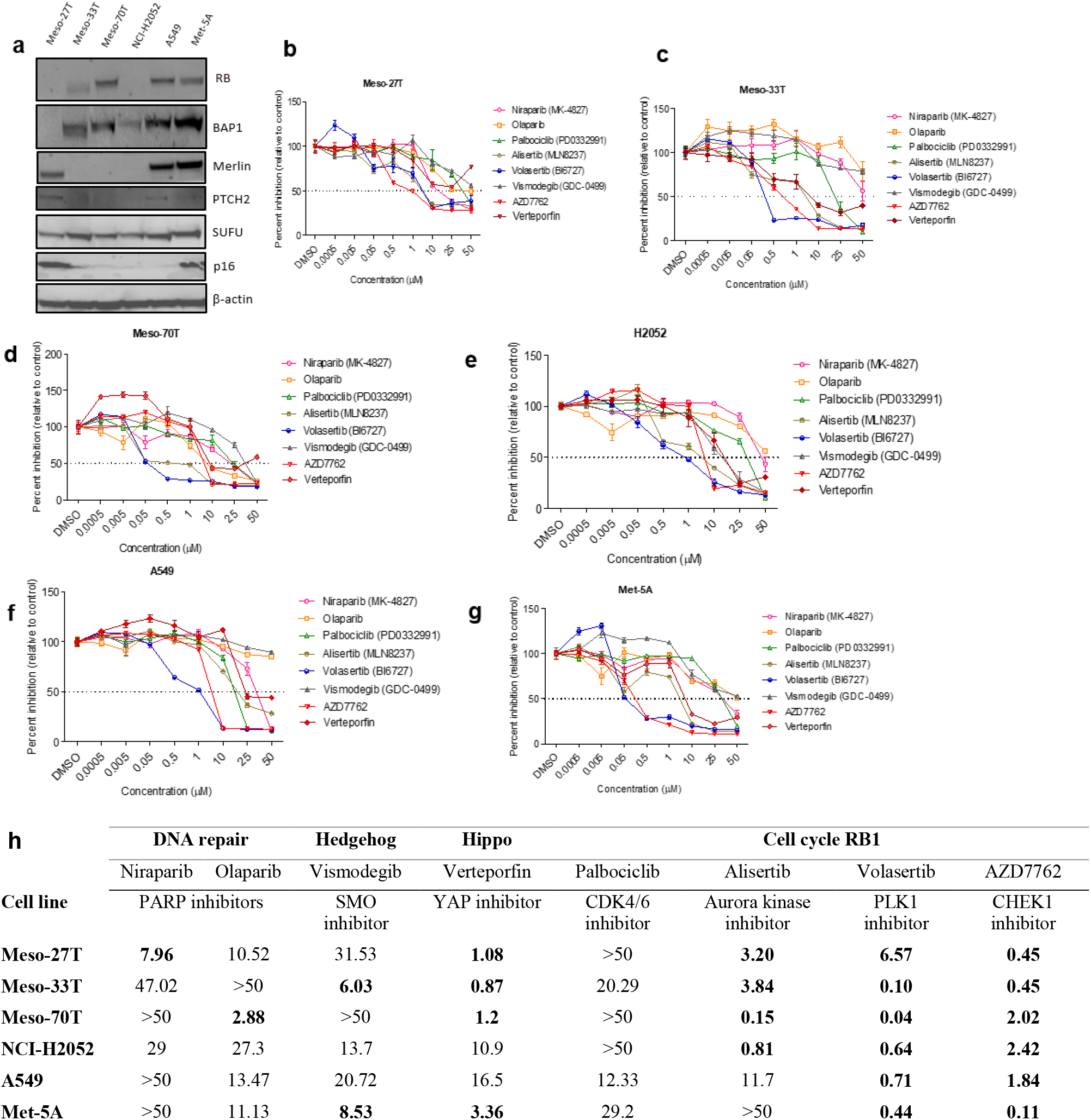
Response of cell lines to tested drugs. a) Characterisation by Western blot of endogenous levels of RB, BAP1, Merlin (NF2), PTCH2, SUFU and p16 (CDKN2A) in primary cells and cell lines (corresponding to mutation/CNA profiles); b-g) Sensitivity dose curves for patient-derived MPM primary cells and cell lines against eight compounds. Standard errors of the mean are shown as bars. Dashed lines mark 50% inhibition; h) IC_50_ values for responses normalized to the control (DMSO), calculated by fitting a dose-response curve model in Graph Pad Prism and tabulated as concentration of drugs in µM. In cases where responses were above the highest drug concentration used in the experiment, IC_50_ estimates are marked as >50µM. Values in bold depict sensitive primary cells and cell lines, where IC_50_ to compounds <10µM and R^2^ (goodness of fit of curve) >0.7

As we had observed mutation signatures associated with repair of DNA double-strand breaks (COSMIC mutational signature 3), we tested two PARP inhibitors (Niraparib and Olaparib) that are effective in homologous repair deficiency^40^. Despite reports suggesting utility in MPM^41^, we did not see a consistent inhibition of primary cell growth (Figure 4h). Our results are supported by a recent report by Fennell *et al*. ^42^ that tested Rucaparib, a third PARP inhibitor, in *BAP1* or *BRCA1* defective MPM patients. Although the clinical trial achieved its endpoints, the author suggested that there is no correlation between BAP1 or *BRCA1* expression and patients’ objective response to Rucaparib.

Another tested drug in our screen, Palbociclib, PD0332991, a CDK4/CDK6 inhibitor, that is supposed to target the most frequent deletion in MPM, *CDKN2A*, showed only minor effects in our cell lines (Figure 4h).

*RB1* loss has recently been shown to confer a robust and selective vulnerability to drugs that target DNA damage checkpoint (*CHEK1*) and chromosome segregation proteins such as Polo-Like-Kinase 1 (*PLK1*)^43^. *CHEK1* is over-expressed in MPM^44^, and RNAi screens have shown MPM lines to be sensitive to *CHEK1* and *PLK1* knockdown. *RB1* deficient tumours are hyper-dependent on Aurora kinase B (AURKB) for survival^45^, and AURKB inhibitors are efficacious against *RB1* deficient lung cancers at non-toxic doses^45^. Consistent with these observations, we found micromolar (µM) to sub-µM responses when treating with an Aurora Kinase inhibitor (Alisertib, MLN8237); an inhibitor of *PLK1/2/3* that induces G2/M arrest and apoptosis (Volasertib, BI6727)^46^; and a *CHEK1/2* inhibitor that abrogates the G2/S checkpoint (AZD7762).

The involvement of Hippo pathways in our results supports previous suggestions that YAP1 axis inhibition may be used in MPM therapy^29^. We found µM IC_50_ responses to the YAP inhibitor Verteporfin in all MPM cell lines (Figure 4h). Notably, immortalised Met-5A mesothelial cells that are not deficient in *NF2* also responded.

## Discussion

In our study we have extended previous genomic analyses by testing copy number aberrations (CNAs) through SNP genotyping arrays together with WES and targeted capture sequencing. We found genetic lesions to be enriched in RB1/cell-cycle, Hippo and Hedgehog pathways, and identified two major immunological influences. We assessed vulnerabilities of MPM tumours by testing drugs that target altered pathways or their members in whole genome sequenced primary MPM cells.

The most frequent genomic alteration in our subjects was deletion of the *CDKN2A* locus on 9p21.3, found in 60% of the analysed samples. This deletion predicts a worse OS than tumours without the deletion^47^. We observed that tumours with this deletion had a higher copy number burden compared with *CDKN2A* wild type patients, consistent with cell cycle dysregulation.

Hippo pathway activation was observed in more than 50% of MPM tumours while Hedgehog pathway activation, as *SUFU* deletion, was seen in 21% of the tumours.

*Cdkn2a* loss and Hedgehog and Hippo pathway activation have been observed in murine models of asbestos exposure well before tumour development^7,8^,19. Our results and the remarkable consistency of genetic lesions in MPM in humans^4,5^ and in mice^7,8^ suggest a hypothesis that recurrent MPM breakpoints and mutations occur in regions of chromatin that have been accessed during the inflammatory response to asbestos. Activation of Hedgehog might also contribute to the stroma-rich microenvironment that characterises MPM tumours.

Our finding of *RB1* deletions in 34% of tumours with *CDKN2A* loss makes responses to CDK4/CDK6 antagonists less likely and underpins our finding that the CDK4/CDK6 inhibitor Palbociclib had marginal effects on primary cell survival. However, we note a previous publication has reported MPM cell lines to be sensitive to this compound^48^ and further investigations are indicated.

We therefore tested compounds downstream of RB1 and showed that *RB1* defective primary cells responded well (irrespective of *CDKN2A* deletion) to an Aurora Kinase inhibitor (Alisertib, MLN8237); an inhibitor of *PLK1/2/3* that induces G2/M arrest and apoptosis (Volasertib, BI6727) and a *CHEK1/2* inhibitor (AZD7762). These findings encourage the clinical investigation of these or related compounds, and a Phase II Trial of Alisertib in salvage malignant mesothelioma is currently under way (NCT02293005).

We identified a recurrent novel amplification of *RASSF7* in 31% of tumours. Taken with other Hippo pathway members (*NF2, LATS1* and *LATS2*), 50% of tumours had at least one lesion of this pathway. Our testing of primary cells revealed micromolar responsiveness of MPM to the YAP inhibitor Verteporfin, although it did not seem to depend on the presence of *NF2* or other Hippo mutations. These results are consistent with recent studies that have shown Verteporfin to be effective *in vitro* against MPM cells^49, 50^.

*BAP1* is the archetypal MPM gene^51^ and was mutated in 31% of tumours and deleted in 33%. We gained some insight into its function by comparing transcriptome abundances between *BAP1* mutation/deletion and *BAP1* wild type tumours, where we found replicated up-regulation of the *RET* proto-oncogene. These results suggest tumour suppressor activities of *BAP1* beyond deubiquitination^52^. It may be of interest that RET inhibitors are effective in RET-driven NCSLC and thyroid cancers^53^.

An important finding of our study was deletion of *SUFU* locus on chromosome 10q24.32 in 21% of tumours. However, we did not find *SUFU* deletions in primary cells and Vismodegib, a Hedgehog inhibitor, was efficient in only one primary cell line. Beneficial effects of Vismodegib on tumour-stromal interactions have previously been shown^27^, and a role for Hedgehog pathways in mesothelial-matrix interactions (as opposed to simple driving of cell division) is further suggested by our findings of the upregulation of *PTCH2, GJB2, NHS* and *HOXA7* in *SUFU* deleted tumours (Table 1). These results encourage the speculation that Vismodegib may be of clinical use to modify MPM fibrosis. Together with *SUFU*, two other genes were deleted at the same locus, *ARL3* and *TRIM8*, the latter being involved in innate immunity.

A striking novel finding in RNA-sequencing expression data of tumours with *SUFU* locus loss was the downregulation of T-cell and antigen-presenting cell genes (Table 1). Although unexpected, these findings were strongly replicated in other data (Supplementary File 1_Table 5) and are consistent with the known central function of Hedgehog signalling in T-cells at the immunological synapse^54,55^.

High levels of VISTA, an alternative checkpoint inhibitor, have previously been reported in MPM and confer a better outcome^4,39^. We confirmed the strong staining for VISTA by IHC and found that *VISTA* abundance strongly correlated with other *SUFU*-affected immune-synapse genes. By contrast, PD-L1 staining was generally weak in the tumours. A small molecule inhibitor against VISTA (CA-170)^56^ is currently in a Phase I clinical trial (NCT02812875), and our findings may provide biomarkers as well as a stimulus further to investigate VISTA therapeutic blockade in MPM.

In the same context, Vismodegib might be considered as an adjuvant to immunotherapy in the presence of *SUFU* loss. It will be relevant to test if aberrant Hedgehog immune signalling is detectable in other malignancies.

It may also be of interest that the Type I Interferon genes on 9p21.3 were deleted in 52% of all MPM. Interferons induce complex pro-inflammatory responses within tumour cells as well as in accessory immune cells^57,58^. Homozygous deletion of *IFN* genes is associated with poor response to CTLA4 blockade in patients with malignant melanoma^59^. Historically, administration of IFNA2 to patients with MPM has occasionally induced complete regression^60,61^. Supplementary, early stage trials suggest that intra-pleural infection with viral vectors containing *IFNA2*^62^ or *IFNB1*^63^ induce inflammation and encourage beneficial MPM responses, suggesting an adjuvant role for interferons in therapy.

The association with *CDKN2A* loss with higher copy number burdens might indicate a beneficial effect of immune checkpoint inhibitors, but the co-deletion of *IFN* type I could enhance tumour cell evasion of immune surveillance. On the other hand, the loss of *IFN* genes may encourage the use of oncolytic virus as therapies^17^. These alternatives could be explored in immunocompetent murine models of MPM.

The results of our investigations should be interpreted in the light of several limitations. We were not powered to investigate determinants of fibrosis, an important variable feature of MPM. Nevertheless, we examined tumours with tumour extent down to 20% as estimated by ASCAT analysis, compared to TCGA analysis of MPM that was confined to tumours with only >70% MPM cells^4^, providing a reference for one extreme of the range.

The dose of asbestos exposure is a known determinant of fibrosis, but not of MPM^64^. Sixty percent of our cases reported working with asbestos, but we did not have accurate estimates of exposure from a detailed occupational history or from fibre counts in unaffected lungs to include exposure as a covariate in our analysis.

Calling the presence of small or uncommon amplifications and deletions in heterogenous tumour samples may be problematic. The amplification segment sizes that we observed in the Infinium SNP array (with ∼3Kb spacing between SNPs) were on average seven times smaller than deletion segments (35.1Kb vs. 265.8Kb), and will have been harder to detect in the uneven coverage of WES. Nevertheless, the novel CNAs are detectable in the large Bueno *et al*. dataset^5^, each showing significant associations with the abundance of their transcribed RNA.

We have shown that genomic findings correlate with protein levels in histological sections of FFPE. Notably, we confirmed a high level of VISTA staining^4,39^ which our transcriptomic data shows to be related to Hedgehog aberration. We were not powered to test systematically for the determinants of histological subtypes or histological features which may be important in clinical decision making. Therefore, there is an unmet need for investigations of the relationship between histology and genomic features identified in this and other studies.

We found distinctive transcriptome changes for some common lesions (*BAP1* and *SUFU* loss), but not for lesions in the RB1 or NF2 pathways. This may be due to lack of power, or possibly to differences in gene expression that result from either acceleration or braking of cellular division.

We tested tool therapeutic compounds as monotherapy in patient-derived low-passage MPM cells. We selected primary cells and cell lines for the drug screen to include the main histological subtypes of MPM and to cover the main drivers of MPM. Although the cell lines contained the most common mutations, some CNAs, such as *RASSF7* amplification were not found. We speculate that they may have been lost by weaning of pure MPM cultures from other cell types. Compounds were chosen for testing to target the pathways shown by our genomic analysis. The known interactions of MPM with stroma suggest better understanding of a wider range of drug effects may come from 3D models that include fibroblasts and immune cells.

## Conclusions

In conclusion, our analyses suggest roles for Aurora Kinase, PLK, CHEK and YAP inhibitors in the treatment of MPM growth. *IFN* Type I and *SUFU* deletions as biomarkers may guide more effective immunotherapies. VISTA inhibition may directly modify immune recognition of MPM, and an adjuvant role in immunotherapy seems possible for Hedgehog inhibitors. The involvement of Hippo and Hedgehog signals and the intense fibrosis seen clinically assert a central role for tumour-matrix interactions in the pathogenesis of MPM and suggest therapeutic avenues beyond tumour cell killing.

## Methods

### Sample collection

Thirty unpaired tumour samples were obtained from the NIHR-BRU Advanced Lung Disease Biobank and Royal Brompton and the Harefield NHS Trust (RBH) Diagnostic Tissue Bank (NRES:10/H0504/9 and 10/H0504/29-NRES Committee Southampton and South West Hampshire) from non-consecutive surgical biopsy samples between 2006 and 2011. Eighty-two paired (tumour and blood) and 4 unpaired tumour samples were obtained from MesobanK UK, Cambridge (NRES:13/EE/0169-East of England - Cambridge Central Research Ethics Committee). Seven additional paired samples were obtained from the EQUALITY study (NRES:10/H0808/53-NRES Committee London-Dulwich). Samples were not collected sequentially or at set times, and the MesobanK specimens came from multiple centres. Tissues were collected during open or thorascopic surgical procedures prior to any anti-cancer treatment and were immediately snap frozen with or without RNAlater. Formalin-fixed paraffin embedded tissues were sectioned, stained with routine haematoxylin and eosin, and reviewed by two experienced pathologists to verify tumour histology and abundance. For RBH and EQUALITY tumours, tissues with 30% or more viable-appearing malignant cells were selected for whole exome sequencing (WES). MesobanK samples were selected to contain only tumours with > 50% malignant cells on histology. Supplementary File 2_Table 1 contains a summary of patients’ clinico-pathologic details. All experiments were performed in accordance with ethical guidelines and all participants gave an informed written consent.

### Genomic DNA isolation and quality control

Genomic DNA was isolated from frozen tumour tissues and matched normal tissue (blood) with routine methods (Qiagen DNA and RNA extraction kits, Qiagen, Hilden, Germany). DNA yield and purity were assessed with the Quant-iT(tm) PicoGreen dsDNA Assay Kit (Life Technologies, Carlsbad, USA) or Qubit 3.0 fluorimeter (Thermo Scientific, Massachusetts, USA) according to manufacturers’ protocols.

### Whole exome sequencing (WES)

WES was performed at the McGill Genome Centre, Canada. Genomic DNA from tumour and blood samples were fragmented and hybridised as per SureSelect^XT^ Target Enrichment System (Agilent SureSelect Human All Exon V4) for the Paired-End Multiplex Sequencing protocol. The captured libraries were sequenced on an Illumina HiSeq2000 according to standard protocols. Supplementary File 2_Table 2 contains a list of somatic variants identified from whole exome sequencing.

### Targeted capture sequencing of a custom gene panel

The entire coding regions of fifty-seven genes were included in a hybridisation capture panel (Supplementary File 1_Table 1), based on: recurrence in our WES tumour set; reported in the Catalogue of Somatic Mutations in Cancer (COSMIC) database; implicated in cancer ^65^; or reported in the TCGA or Bueno *et al*. studies^4,5^. Sequencing libraries were prepared from DNA extracted from tumours and normal tissue (whole blood) samples using the SureSelect QXT Target Enrichment System (Agilent, Santa Clara, USA) according to the manufacturer’s protocols. Sequencing was performed on a MiSeq or NextSeq500/550 platform (Illumina) with a mean read depth of 780.6X (all samples). Supplementary File 2_Table 3 contains a list of filtered somatic variants from targeted capture sequencing.

### Data processing and quality control

Raw fastq files were quality checked before alignment with BWA mem (v 0.7.12). GATK software (v 3.8 and 4.1) was used to refine the alignment data before variant calling. For Target capture sequencing (TCS), somatic and germline variant calling was performed for the paired samples using VarScan software (v 2.4.2). For the un-paired samples joint variant calling was performed using Platypus (v 0.8.1). In case of WES, joint variant calling was performed at McGill using the GATK HaplotypeCaller.

### Detection of candidate pathogenic somatic and germline variants

Candidate somatic and germline variants were checked for presence in population data and those with frequency >=10^−3^ were to be polymorphisms and filtered out. Further selection was based on either being assigned as High or Moderate impact by VEP^66^ or predicted to be splice-site altering (dbscSNV^67^ score of >0.6) by at least two of three algorithms. Further prioritisation of SNV (single nucleotide variant) candidates was done based on predicted deleteriousness from any one of SIFT, Polyphen and MutationTaster algorithms.

### Copy-number analysis

One hundred and twenty-one DNA samples were interrogated at Eurofins against the Human Infinium Omni-Express-Exome v 1.3 and v 1.4 Bead Chips (Illumina) arrays containing >950K markers. One hundred and eighteen samples remained (77 paired and 43 unpaired) after QC checks. Raw copy number data (LRR and BAF) were exported from GenomeStudio software (v 1.9.4). GC correction was performed to account for genomic wave artefacts affecting SNP arrays using ASCAT (v 2.4.4). The GC corrected Log R ratios (LRR) were then processed using DNACopy (v 1.52) for segmentation and filtered for marker support. Recurrent germline CN segments were identified and subtracted from the tumour sample CN segments. Germline subtracted copy number segments were then processed with GISTIC (v 2.0.23). Plotting of GISTIC results was done in maftools (v 1.4.28). Copy number burden was defined as the percentage of the genome carrying copy number aberration.

### Mutation signature analysis

Somatic single nucleotide variants (SNVs) from the 21 paired WES samples were analysed for tri-nucleotide frequency around the mutated base using MutationalPatterns^68^ in R. The Sanger COSMIC signature panel (n=30) was used to infer mutational processes by obtaining the percentage contribution of 30 signatures per sample. Only signatures contributing to >25% of samples were carried forward. Paired germline samples were not available for whole-genome sequencing (WGS) of 19 MPM primary cells (PMCC) (one MPM primary cell WGS had failed QC), and so annotation-assisted filtering of the total SNVs was done. Only those SNVs that were non-polymorphic and either protein-sequence altering or predicted to be splice-site altering, were considered and analysed as described for the tumour tissue samples (Supplementary File 1_Figure 6).

### RNA sequencing

Total RNA was isolated from 35 tumours using the RNEasy Fibrous Midi kit (Qiagen, Hilden, Germany) following the manufacturer’s protocol. Concentration and quality were determined with the 2100 Bioanalyzer and Total RNA Nano kit (Agilent Technologies, California, United States) as per manufacturer’s instructions. RNA sequencing was performed at McGill Genome Centre.

### Replication in published data

A Supplementary panel of 99 paired mesothelioma samples analysed by whole-exome sequencing (WES) and published by Bueno *et al*. ^5^ was downloaded to investigate the presence of CNAs for *RASSF7, RB1* and *SUFU*. The concurrent RNA-sequencing (RNA-seq) (98 of the WES samples) was also investigated for expression change in the direction of copy-number. Raw fastq reads were accessed from the EGA repository (EGAS00001001563). After quality-checking sequences were aligned using BWA MEM^69^ and STAR^70^ for WES and RNA-seq data respectively. CNVkit^71^ was used to estimate CN calls. Mann-Whitney tests were used for comparisons between CNA versus CNA neutral samples.

### Immunohistochemistry

Three μm whole slide FFPE tumours sections mirroring fresh frozen tissue used for molecular analysis underwent H&E staining according to routine histopathological protocols. Further sections underwent staining for BAP-1 (Santa Cruz BioTechnology, clone C4), Ki67 (Ventana, 30-9), MTAP (NovusBio, 2G4), PD-L1 (Ventana, SP263) and VISTA (D1L2G, Cell Signalling Technology). Mitotic activity was evaluated by counting the number of mitotic figures in the area of highest activity, over 10 high powered fields (0.24mm^2^).

### Whole genome sequencing of primary cell lines

Genomic DNA extracted from patient derived MPM cell lines (n=20, of which one failed QC) and primary normal mesothelial cells, MES-F (purchased from ZenBio, USA) underwent WGS (McGill) and SNP genotyping (Eurofins). Genomic details of the commercial cell line were obtained from published data (COSMIC, CCLE databases, 61). Supplementary File 2_Table 4 contains a list of variants from whole genome sequencing for the primary cells.

### *In vitro* drug testing

Patient-derived primary cells, Meso-27T, Meso-33T and Meso-70T were obtained from the MRC Toxicology Unit, University of Cambridge, UK. Commercial cell lines NCI-H2052 (sarcomatoid mesothelioma), A549 (lung adenocarcinoma) and Met-5A (normal mesothelial, SV40 transformed) previously obtained from ATCC were gifted from the MRC Toxicology Unit. Original establishment of the primary cells was as previously described^12^. All primary cells and cell lines were maintained in RPMI-1640 growth media supplemented with L-glutamine (2 mM), penicillin (100 U/ml), streptomycin (100 μg/ml) and 10% FBS at 37 °C in 5% CO_2_.

Eight drugs were investigated, based on our results. These were Niraparib (MK-4827, HY-10619, MedChem Express), Olaparib (HY-10162, MedChem Express), Palbociclib, PD0332991 (A8316, ApexBio), Alisertib (MLN8237, S1133, Selleckchem), Volasertib (BI6727, S2235, Selleckcehm), Vismodegib (GDC-0499, S1082, Selleckchem), AZD7762 (S1532, Selleckchem) and Verteporfin (SML0534, Sigma Aldrich). All drugs were diluted in DMSO and aliquots maintained at −-20°C. Drug aliquots were freeze-thawed no more than three times. For all experiments, controls consisted of DMSO-alone treated primary cells or cell lines.

Cells were seeded in 96-well plates (4 x 10^3^ cells/well) 24h prior to drug treatments. Each line was treated for six days (except for drug PD0332991 where treatment was 3 days) with a range of concentrations from 0.0005 to 50μM. Cell viability was measured with MTS assay (CellTiter 96® AQueous One Solution Cell Proliferation Assay, Promega) on a plate reader (Tecan).

Three independent experiments, each having three technical replicates, were conducted for each drug tested. Results are represented as the average normalized to the control at each time point (mean ± s.e.m.). Briefly, the raw optical densities obtained from each well were normalized to the average of DMSO control wells, that was considered 100% viability (maximal DMSO concentration used was 0.5%). IC_50_ values were calculated with Graph Pad Prism 5 software (GraphPad Software Inc, San Diego, CA) using a dose-response curve fit model using the nonlinear log (inhibitor) versus response-variable slope (four parameters) equation. In addition, the IC_50_ values were only considered if the software gave unambiguous results and the R^2^ value was > 0.7.

### Statistical analysis

Categorical variables were evaluated using Fisher’s exact test for two-by-two comparison or Pearson’s χ^2^ for comparison that exceeded the two-by-two condition. Differences between groups were evaluated by means of nonparametric Mann-Whitney or Kruskal-Wallis test.

### Clinical outcomes

Overall survival (OS), defined as time from date of diagnosis to time of death, was available for 110 patients. OS was estimated using the Cox-Mantel log-rank test, Kaplan-Meier method. Censoring of OS was done at the date of the last follow-up if death did not occur. Survival analyses were performed using the long-rank Kaplan-Meier and the differences in survival curves were assessed by Mantel Cox Log rank test. A *P*≤0.05 was considered statistically significant and noted as: \**P*≤0.05, \*\**P*≤0.01, \*\*\**P*≤0.001. Tests and graphs were performed with Graph-Pad Prism 5, SPSS Statistics 25 or R Studio.

## Data Availability

All sequence data is being depositited with the European Genome Archive and will be made freely available following acceptance in a peer-reviewed journal

## Acknowledgements (Funding)

This study was financially supported by a Libor Fund grant from the UK Department of Health, by the British Lung Foundation and by the Asmarley Foundation. MMF, AEW, XMS and TC are funded by the UK Medical Research Council. SP acknowledges NHS funding to the Royal Marsden Hospital-Institute of Cancer Research Biomedical Research Centre. We thank Cambridge Biomedical Research Centre and Cambridge Cancer Centre.

## Declarations

### Authors’ contributions

**WOCMC, MFM, ML, SP** and **AMB** planned the study; **AN** processed samples and prepared libraries for targeted capture sequencing, post-analysed whole exome, TCS-NGS, RNA sequencing and IHC data, and performed drug testing on primary cells and cell lines; **AM** performed bioinformatics analysis of whole exome and whole-genome sequencing, target capture, RNA sequencing and copy number data; **SG** and **SKL** processed and prepared samples for whole exome, RNA sequencing and copy number analysis; **HA** analysed WES data; **DMR** and **ME** provided support in generating the TCS-NGS panel and analysis of the libraries; **AB** performed immunohistochemistry staining, **YZZ** and **AGN** interpreted immunohistochemistry data; **TC, XMS, AEW** and **MMF** established and characterized primary cells from tumours; **ML** oversaw whole-exome and RNA sequencing of the samples and advised on analyses; **EL, RCR** and **TB** gathered samples and associated metadata for the study; and **SP** and **ANT** provided clinical and epidemiological scientific guidance. **AN** and **AM** wrote the first draft of the manuscript with guidance by **WOCMC, MFM** and **AMB**, before editing by all the other authors.

### Ethics approval and consent to participate

All samples were collected and used under ethic and consented approval to participate in the study. Thirty unpaired tumour samples were obtained from the NIHR-BRU Advanced Lung Disease Biobank and Royal Brompton and the Harefield NHS Trust (RBH) Diagnostic Tissue Bank (NRES:10/H0504/9 and 10/H0504/29-NRES Committee Southampton and South West Hampshire) with eighty-two paired (tumour and blood) and 4 unpaired tumour samples obtained from MesobanK UK, Cambridge (NRES:13/EE/0169-East of England - Cambridge Central Research Ethics Committee). Seven Supplementary paired samples were obtained from the EQUALITY study (NRES:10/H0808/53-NRES Committee London-Dulwich).

### Consent for publication

Non applicable

### Availability of data and materials

The datasets generated and/or analysed during the current study have been submitted at the European Genome-Phenome Archive (EGA) (https://ega-archive.org/) under accession code: EGAS00001004845.

