## Supplementary File 1 for "Integrated genomics identify novel immunotherapy targets for malignant mesothelioma"

SUPPLEMENTARY DATA

Anca Nastase[1¶], Amit Mandal[1¶], Shir Kiong Lu[1], Hima Anbunathan[1], Deborah Morris-Rosendahl[1,2], Yu Zhi Zhang[1,3], Xiao-Ming Sun[4], Spyridon Gennatas[1], Robert C Rintoul[5,6], Matthew Edwards[2], Alex Bowman[3], Tatyana Chernova[4], Tim Benepal[7], Eric Lim[8], Anthony Newman Taylor[1], Andrew G Nicholson[1,3], Sanjay Popat[9,10], Anne E Willis[4], Marion MacFarlane[4], Mark Lathrop[11], Anne M Bowcock[1], Miriam F Moffatt[1,*], William OCM Cookson[1,*]

Affiliations

1 National Centre for Mesothelioma Research, Imperial College London, UK

2 Clinical Genetics and Genomics, Royal Brompton and Harefield NHS Foundation Trust, London, UK

3 Department of Histopathology, Royal Brompton and Harefield NHS Foundation Trust, London, UK

4 Medical Research Council Toxicology Unit, University of Cambridge, Cambridge, UK

5 Department of Thoracic Oncology, Papworth Hospital, Cambridge, UK

6 Department of Oncology, University of Cambridge, Cambridge, UK

7 Department of Oncology, St George's Healthcare NHS Foundation Trust, London, UK

8 Department of Thoracic Surgery, Royal Brompton and Harefield Foundation Trust, London, UK

9 Department of Medicine, Royal Marsden Hospital, London, UK

10 The Institute of Cancer Research, London, UK

11 Department of Human Genetics, McGill Génome Centre, Québec, Canada

¶ Contributed equally to this study *Contributed equally to this study

*Corresponding authors: Professor William OCM Cookson: National Heart and Lung Institute, Dovehouse Street, London SW36LY, +447788628503; Professor Miriam F Moffatt:

Supplementary_Table 1. Targeted capture next-generation sequencing gene panel

| | | | | | | | |
|---|---|---|---|---|---|---|---|
| *ANKLE1* | *CDKN2A* | *FOXD1* | *MAML3* | *NF2* | *SETD1B* | *TJP2* | *ZNF880* |
| *APC* | *CFAP45* | *HERC1* | *MLH1* | *NRAS* | *SETD2* | *TJP3* | |
| *ASS1* | *CREBBP* | *HERC2* | *MMP17* | *PIK3CA* | *SETD6* | *TP53* | |
| *ATXN2* | *DDX3X* | *ICA1* | *MSH2* | *PRDM12* | *SETDB1* | *TRAF7* | |
| *BAP1* | *DDX51* | *INADL* | *MSH3* | *PRKRA* | *SF3B1* | *ULK2* | |
| *BRCA2* | *EGFR* | *LATS1* | *MSH5* | *PTEN* | *SLC2A3* | *VEZF1* | |
| *BRD4* | *EP400* | *LATS2* | *MSH6* | *RB1* | *SYNE1* | *WDR89* | |
| *CD163* | *FBXW7* | *MACF1* | *NCOR2* | *RYR2* | *TET1* | *ZNF77* | |

### Supplementary_Table 2. Patient demographics

| | Group | | | | | |
|---|---|---|---|---|---|---|
| | **Whole Exome Sequencing (n=50)** | **SNP genotyping (n=118)** | **Targeted Capture Sequencing (n=119)** | **RNA sequencing (n=35)** | **IHC (n=28)** | **All (n=121)** |
| **Gender (n, %)** | | | | | | |
| M | 39 (78.0) | 103 (87.3) | 103 (86.6) | 25 (75.8) | 23 (82.1) | 105 (86.8) |
| F | 11 (22.0) | 15 (12.7) | 16 (13.4) | 8 (24.2) | 5 (17.9) | 16 (13.2) |
| M:F | 3.5:1 | 6.9:1 | 6.4:1 | 3.13:1 | 4.6:1 | 6.6:1 |
| **Age** | | | | | | |
| Average ± SD | 69.7 ± 8.2 | 72.1 ±8.5 | 72.1 ± 8.5 | 70.06 ± 8.3 | 67.6 ± 8.1 | 72.1 ± 8.4 |
| median [range] | 69.7 [52-90] | 73.0 [51-90] | 72.6 [51-90] | 69.0 [55-90] | 67 [52-82] | 73 [51-90] |
| **Histology (n, %)** | | | | | | |
| Epithelioid | 34 (68.0) | 89 (75.4) | 88 (74.0) | 26 (74.3) | 19 (67.9) | 90 (74.4) |
| Biphasic | 14 (28.0) | 23 (19.5) | 25 (21.0) | 6 (17.1) | 9 (32.1) | 25 (20.6) |
| Sarcomatoid | 2 (4.0) | 6 (5.1) | 6 (5.0) | 3 (8.6) | 0 (0.0) | 6 (5.0) |
| **Overall survival** | | | | | | |
| Data available | 92.0% | 91.5% | 92.4% | 85.7% | 100.0% | |
| All (months, [rate at 12 months]) | 9.93 [42.63%] | 9.93 [42.24%] | 9.93 [42.39%] | 10.43 [43.75%] | 12.8 [59.74%] | 9.93 [42.0%] |
| Epithelioid | 10.43 [45.16%] | 11.6 [48.75%] | 11.6 [48.75%] | 12.27 [52.17%] | 12.96 [68.42] | 11.6 [48.15%] |
| Biphasic | 10.86 [42.73%] | 7.4 [29.54%] | 7.4 [31.41%] | 7.07 [33.33%] | 10.87 [38.1%] | 7.4 [31.41%] |
| Sarcomatoid | 1.5 [0.0%] | 3.66 [0.0%] | 3.66 [0.0%] | 1.5 [0.0%] | NA | 3.67 [0.0%] |
| **Asbestos exposure (n, %)** | | | | | | |
| Yes | 32 (64.0) | 63 (68.5) | 63 (67.7) | 20 (57.1) | 21 (75.0) | 83 (68.6) |
| No | 16 (32.0) | 27 (29.3) | 28 (30.1) | 11 (31.4) | 6 (21.4) | 36 (29.8) |
| Unknown | 2 (4.0) | 2 (2.2) | 2 (2.2) | 4 (11.4) | 1 (3.6) | 2 (1.6) |

### Supplementary_Table 3. Significant copy number deletions and amplifications

***DELETIONS***

| cytoband | q value | residual q | wide peak boundaries | Kb | Principal Genes |
|---|---|---|---|---|---|
| **9p21.3** | 2.4E-112 | 2.4E-112 | chr9:21927328-22031004 | 104 | *CDKN2A, CDKN2B, C9orf53* |
| **3p21.1** | 7.3E-10 | 7.3E-10 | chr3:52433746-52449046 | 15 | *BAP1* |
| **10q23.31** | 8.6E-05 | 1.6E-04 | chr10:91440057-91589280 | 149 | *KIF20B, FLJ37201* |
| **16p13.3** | 1.6E-04 | 1.4E-03 | chr16:5128784-7773249 | 2644 | *RBFOX1, FAM86A* |
| **4q13.3** | 2.2E-03 | 3.2E-03 | chr4:70932306-71062425 | 130 | *CSN1S2AP, C4orf40, CSN1S2BP* |
| **6q14.2** | 5.8E-03 | 5.5E-03 | chr6:84563880-84759646 | 196 | *CYB5R4* |
| **11p15.5** | 1.6E-02 | 1.6E-02 | chr11:906709-1083236 | 177 | *AP2A2, MUC6* |
| **10q24.32** | 2.5E-03 | 2.7E-02 | chr10:104245589-104485300 | 240 | *ARL3, SUFU, TRIM8* |
| **1p22.1** | 7.5E-03 | 3.9E-02 | chr1:93620394-93787866 | 167 | *CCDC18* |
| **13q14.2** | 4.2E-02 | 4.2E-02 | chr13:48835311-49063155 | 228 | *RB1, LPAR6* |
| **11q23.2** | 4.7E-02 | 4.9E-02 | chr11:113236976-113280927 | 44 | *ANKK1* |
| **14q22.3** | 4.8E-02 | 4.9E-02 | chr14:56023823-56271005 | 247 | *KTN1, RPL13AP3, LINC00520, KTN1-AS1* |

***AMPLIFICATIONS***

| cytoband | q value | residual q | wide peak boundaries | Kb | Principal Genes |
|---|---|---|---|---|---|
| **11p15.5** | 6.4E-48 | 6.4E-48 | chr11:552679-587371 | 35 | *RASSF7, PHRF1, LRRC56, LOC143666, C11orf35, MIR210, MIR210HG* |
| **19q13.43** | 6.1E-45 | 6.1E-45 | chr19:56572845-56674547 | 102 | *ZNF444, NLRP5, ZNF787* |
| **5q35.2** | 3.8E-39 | 1.2E-37 | chr5:176025071-176043332 | 18 | *GPRIN1, CDHR2* |
| **5q35.3** | 1.8E-10 | 8.3E-08 | chr5:179214819-179249072 | 34 | *LTC4S, SQSTM1, MGAT4B, MIR1229* |
| **2p24.1** | 9.1E-06 | 9.1E-06 | chr2:21210344-21227220 | 17 | *APOB* |
| **19p13.11** | 9.1E-06 | 9.1E-06 | chr19:17339697-17370189 | 30 | *NR2F6, OCEL1, USHBP1* |
| **7p22.3** | 1.2E-04 | 1.2E-04 | chr7:2552807-2579466 | 27 | *LFNG, BRAT1, MIR4648* |
| **16p13.3** | 1.2E-04 | 1.2E-04 | chr16:772127-786320 | 14 | *NARFL, FAM173A, HAGHL, CCDC78* |
| **2q36.3** | 1.6E-04 | 1.6E-04 | chr2:227646960-227686906 | 40 | *IRS1* |
| **16q22.1** | 2.7E-04 | 2.7E-04 | chr16:67220742-67229485 | 9 | *E2F4, EXOC3L1* |
| **6p22.1** | 6.9E-04 | 6.9E-04 | chr6:29311440-29325601 | 14 | *OR5V1* |
| **5q13.2** | 1.5E-06 | 4.7E-03 | chr5:70806522-70855825 | 49 | *BDP1* |
| **11q21** | 1.8E-02 | 1.8E-02 | chr11:95569326-95603081 | 34 | *MTMR2* |
| **6q23.3** | 2.2E-02 | 2.2E-02 | chr6:136582550-136648626 | 66 | *BCLAF1* |
| **19p13.3** | 2.2E-02 | 2.2E-02 | chr19:1457003-1482901 | 26 | *APC2, PCSK4, C19orf25* |
| **16q24.2** | 4.6E-02 | 4.6E-02 | chr16:88498491-89347394 | 849 | *APRT, CBFA2T3, CDH15, CYBA, GALNS, MVD, PIEZO1, IL17C, ANKRD11, TRAPPC2L, CDT1, ZNF469, RNF166, ZC3H18, SLC22A31, ZFPM1, MGC23284, ZNF778, ACSF3, LINC00304, SNAI3, CTU2, PABPN1L, LOC400558, MIR4722* |

### Supplementary_Table 4. Genes differentially regulated in histological subtypes

| Log2 Fold Change | Average Expression | Adjusted P Value | Chr | Name | Description |
|---|---|---|---|---|---|
| **5.23** | -2.62 | 2.10E-04 | 7 | *NFE4* | Nuclear factor in erythrogenesis |
| **4.72** | -0.21 | 1.90E-04 | 3 | *RP4-555D20.2* | miRNA |
| **4.65** | -1.60 | 9.13E-04 | 17 | *IGF2BP1* | Novel interacting partner of p38 MAPK. RNA-binding protein, involved in tumour progression |
| **4.50** | -0.31 | 9.13E-04 | 5 | *GDNF* | Glial cell derived neurotrophic factor |
| **3.66** | 1.57 | 9.92E-04 | 18 | *CCBE1* | High levels contribute to aggressiveness and poor prognosis of Colon Cancer |
| **3.57** | 3.41 | 7.26E-04 | 12 | *WNT5B* | Activator of WNT signalling |
| **3.45** | -2.82 | 9.13E-04 | 15 | *CTD-2033D15.2* | Non-coding cDNA |
| **3.33** | -3.95 | 9.91E-04 | 3 | *RP4-555D20.4* | miRNA |
| **2.89** | -3.88 | 7.18E-04 | X | *RP11-320G24.1* | miRNA |
| **2.66** | 1.29 | 7.26E-04 | 17 | *HS3ST3A1* | Tumour regulator and prognostic marker in breast cancer. |
| **2.27** | 5.55 | 2.10E-04 | 1 | *NAV1* | Potentiates migration of breast cancer cells |
| **2.25** | 3.73 | 7.26E-04 | 2 | *CHN1* | Actin dynamics in cell migration |
| **2.15** | 3.41 | 7.26E-04 | 15 | *GPR176* | Orphan G-protein-coupled receptor that sets the pace of circadian behaviour |
| **1.95** | 6.25 | 2.58E-04 | 4 | *SEPT11* | |
| **1.73** | 5.81 | 9.57E-04 | 6 | *TRAM2* | Putative metastatic factor for oral cancer |
| **1.50** | 3.88 | 9.57E-04 | 2 | *SERTAD2* | Promotes oncogenesis in nude mice and is frequently overexpressed in multiple human tumours. |

Genes differentially upregulated in sarcomatoid and mixed histology compared to epithelioid. Transcripts with average expression ≥ 1 are shown. *P* values are adjusted for multiple comparisons (false discovery rate <0.05).

### Supplementary_Table 5. Correlation of *SUFU* and immune function genes and replication

| NCMR n=35 | ***SUFU*** | | ***PTCH2*** | | ***PTCH1*** | | ***CR1*** | | ***KLRD1*** | | ***PD-L1*** | |
|---|---|---|---|---|---|---|---|---|---|---|---|---|
| ***PTCH2*** | -0.38 | *2.5E-02* | | | | | | | | | | |
| ***PTCH1*** | -0.37 | *3.1E-02* | 0.75 | *1.7E-07* | | | | | | | | |
| ***CR1*** | 0.50 | *2.2E-03* | -0.59 | *1.8E-04* | -0.41 | *1.4E-02* | | | | | | |
| ***KLRD1*** | 0.37 | *2.9E-02* | -0.60 | *1.4E-04* | -0.52 | *1.3E-03* | 0.56 | *5.1E-04* | | | | |
| **PD-L1** | 0.10 | *5.8E-01* | -0.39 | *2.2E-02* | -0.44 | *8.1E-03* | 0.35 | *4.1E-02* | 0.37 | *2.8E-02* | | |
| **VISTA** | 0.61 | *1.1E-04* | -0.52 | *1.3E-03* | -0.41 | *1.3E-02* | 0.43 | *1.0E-02* | 0.68 | *7.5E-06* | 0.09 | *6.1E-01* |
| TGCA-Meso n=86 | | | | | | | | | | | | |
| ***PTCH2*** | -0.08 | *4.6E-01* | | | | | | | | | | |
| ***PTCH1*** | 0.09 | *3.9E-01* | 0.77 | *2.2E-16* | | | | | | | | |
| ***CR1*** | -0.04 | *7.1E-01* | -0.09 | *4.1E-01* | -0.11 | *3.4E-01* | | | | | | |
| ***KLRD1*** | -0.04 | *7.0E-01* | -0.36 | *6.2E-04* | -0.33 | *1.9E-03* | 0.41 | *7.9E-05* | | | | |
| **PD-L1** | -0.26 | *1.7E-02* | -0.22 | *4.0E-02* | -0.30 | *5.5E-03* | 0.35 | *8.1E-04* | 0.40 | *1.7E-04* | | |
| **VISTA** | 0.25 | *2.1E-02* | -0.47 | *5.9E-06* | -0.25 | *2.2E-02* | -0.06 | *5.9E-01* | 0.37 | *4.8E-04* | -0.09 | *4.0E-01* |
| Bueno *et al*. n=211 | | | | | | | | | | | | |
| ***PTCH2*** | -0.22 | *1.4E-03* | | | | | | | | | | |
| ***PTCH1*** | -0.16 | *1.8E-02* | 0.71 | *2.2E-16* | | | | | | | | |
| ***CR1*** | 0.30 | *9.6E-06* | -0.20 | *2.9E-03* | -0.23 | *7.9E-04* | | | | | | |
| ***KLRD1*** | 0.26 | *1.1E-04* | -0.37 | *3.1E-08* | -0.35 | *2.4E-07* | 0.49 | *7.1E-14* | | | | |
| **PD-L1** | -0.08 | *2.3E-01* | -0.16 | *1.7E-02* | -0.31 | *6.0E-06* | 0.42 | *2.8E-10* | 0.38 | *1.2E-08* | | |
| **VISTA** | 0.27 | *5.6E-05* | -0.36 | *6.0E-08* | -0.27 | *8.3E-05* | 0.06 | *3.9E-01* | 0.36 | *6.8E-08* | -0.06 | *3.5E-01* |
| Combined studies | | | | | | | | | | | | |
| ***PTCH2*** | -0.21 | *9.96E-05* | | | | | | | | | | |
| ***PTCH1*** | -0.14 | *1.20E-02* | 0.73 | *<2.2E-16* | | | | | | | | |
| ***CR1*** | 0.16 | *3.53E-03* | -0.17 | *2.23E-03* | -0.15 | *6.87E-03* | | | | | | |
| ***KLRD1*** | 0.14 | *9.81E-03* | -0.33 | *6.71E-10* | -0.28 | *3.33E-07* | 0.55 | *<2.2E-16* | | | | |
| **PD-L1** | -0.09 | *8.96E-02* | -0.18 | *7.87E-04* | -0.28 | *3.31E-07* | 0.42 | *1.78E-15* | 0.42 | *1.33E-15* | | |
| **VISTA** | 0.33 | *6.93E-10* | -0.42 | *2.00E-15* | -0.32 | *3.81E-09* | -0.06 | *2.66E-01* | 0.22 | *5.59E-05* | -0.11 | *5.50E-02* |

Pearson correlations between abundances of Hedgehog pathway transcripts *SUFU*, *PTCH1*, *PTCH2* and transcripts related to immune checkpoints. The official gene names for PD-L1 and VISTA are *CD274* and *VSIR*, respectively. Results are shown for the present study, two previous investigations and for all studies combined. Two sided-*P* values are shown in italics throughout.

Supplementary_Table 6. Genes differentially up-regulated in *BAP1* mutated tumours

| Log2 Fold Change | Average Expression | Adjusted P Value | Chr | Name | Description |
|---|---|---|---|---|---|
| **6.09** | 4.33 | 1.88E-02 | 20 | *NNAT* | Marker of poor outcome in many cancers |
| **3.60** | 1.47 | 7.77E-03 | 16 | *NECAB2* | Neuronal calcium-binding protein that binds to adenosine A(2A) and metabotropic glutamate type 5 receptors |
| **3.20** | 3.51 | 4.77E-02 | 22 | *TAFA5* | Postulated to function as brain-specific chemokine or neurokine |
| **2.70** | 2.43 | 1.88E-02 | 21 | *PDE9A* | Phosphodiesterase 9A |
| **2.36** | 1.68 | 4.77E-02 | 10 | *RET* | Tyrosine kinase receptor leading to oncogenic signalling that is targetable with anti-RET multikinase inhibitors |
| **1.85** | 4.08 | 4.77E-02 | 19 | *HSD17B14* | Hydroxysteroid 17-beta dehydrogenase 14 |

### Supplementary_Figure 1. Analytical structure

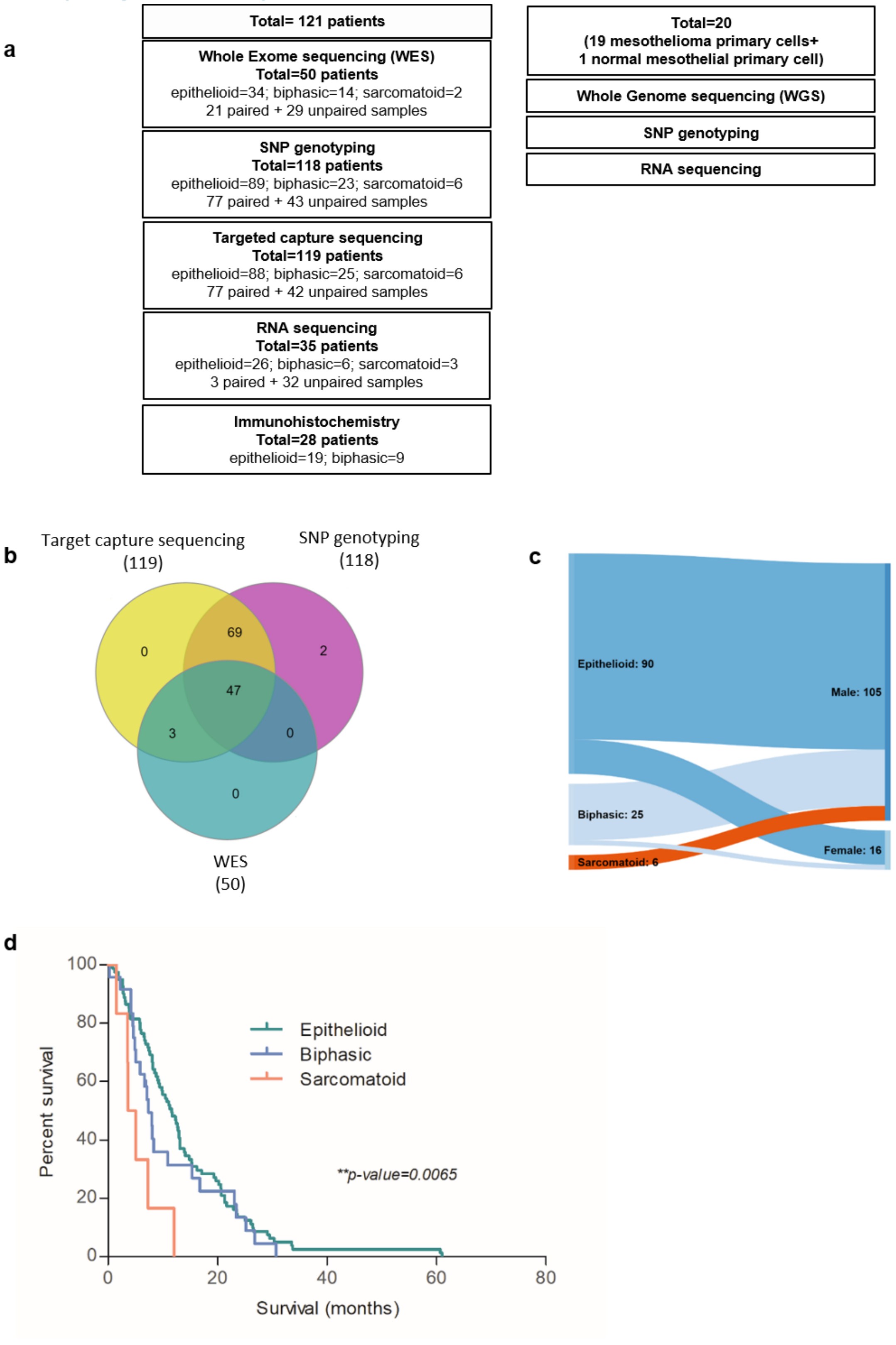

**a**) Summary of samples used in each analysis**; b)** Venn diagram showing overlapping of samples used in WES, SNP genotyping and targeted capture sequencing (please refer to Supplementary File 2- Table1 for further details); **c**) Sankey diagram on histological subtype and patients` gender; **d**) Kaplan-Meier survival curves on histological subtypes

### Supplementary_Figure 2. Oncoplot from targeted sequencing panel

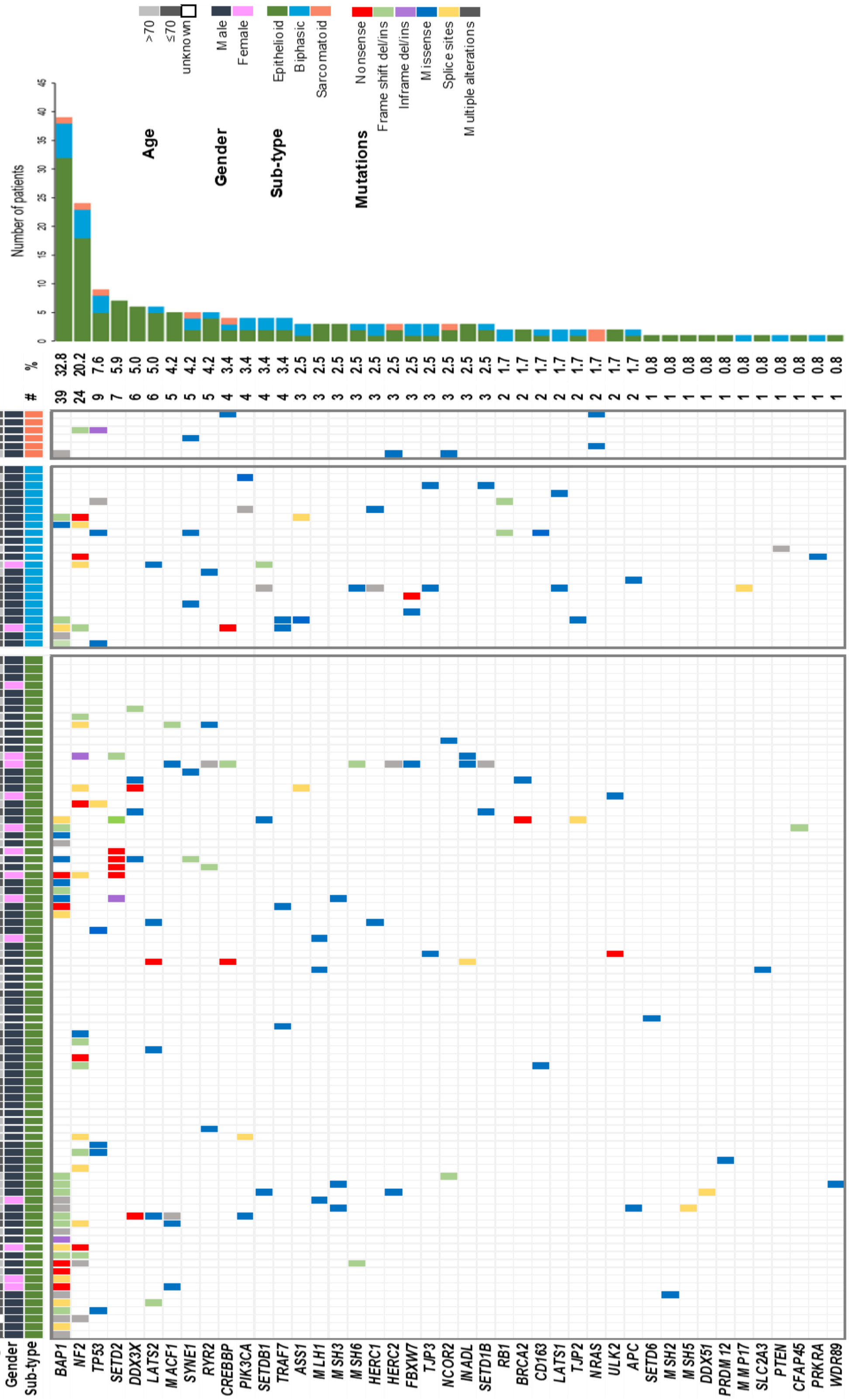

Oncoplot summarizing results from the targeted capture panel analysis. Genes are arranged down from the most frequently mutated Three panels are shown, each panel representing a histologic subtype (epithelioid, biphasic and sarcomatoid) with additional information related to age and gender. The most frequent mutations for each gene, coloured by subtype are shown as a bar chart.

### Supplementary_Figure 3. Mutation characteristics

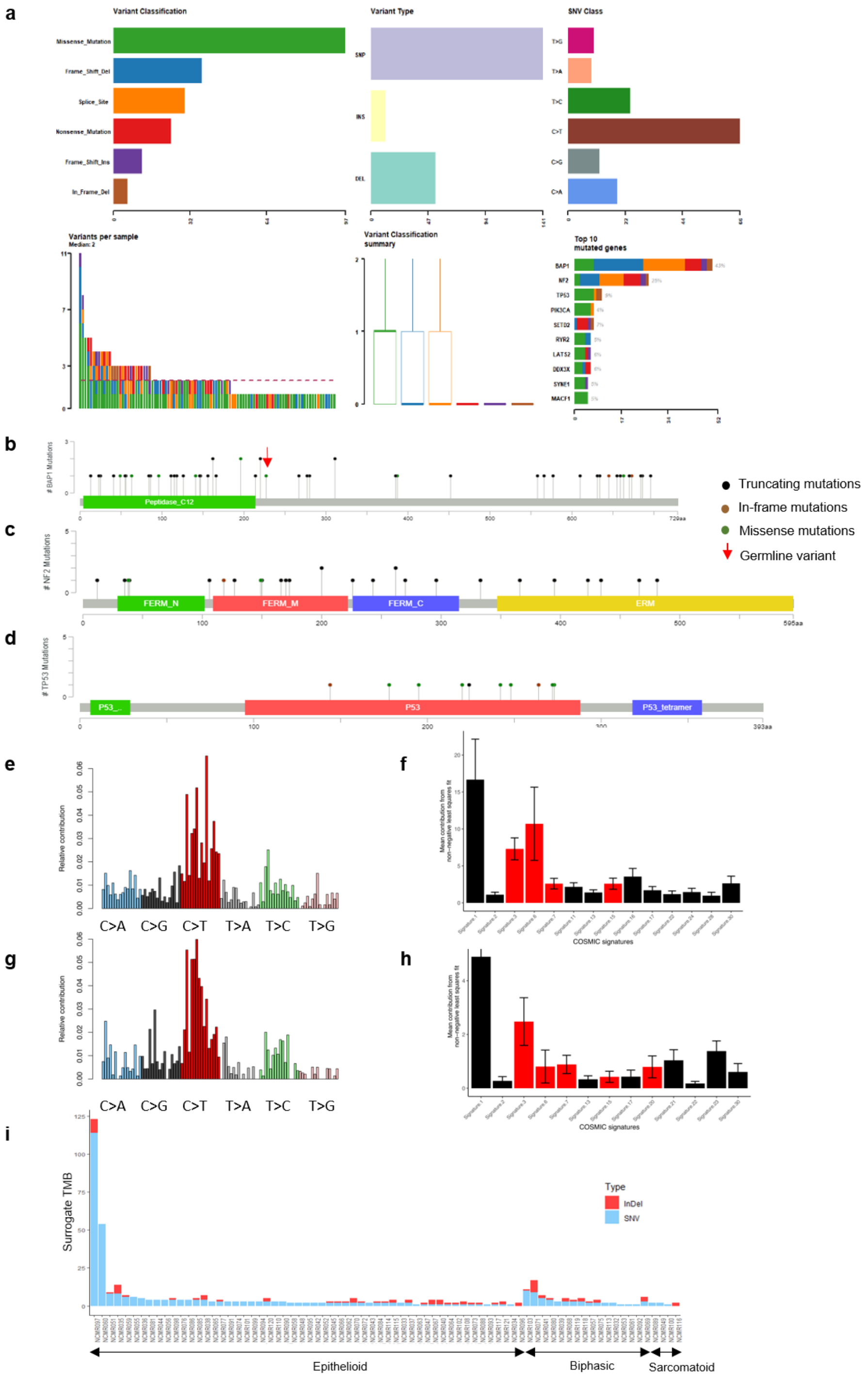

**a**) Summary of mutation spectrum observed for genes from targeted capture sequencing panel; **b-d**) Distribution of mutations in *BAP1*, *NF2*, and *TP53*; **e**) Mutational signature in 21 paired MPM analysed by WES; **f**) Mean percentage of contribution for COSMIC signatures: red bars indicate signatures associated with DNA damage; **g**) Mutational signature in 19 patient-derived MPM cell lines; **h**) Mean percentage COSMIC signatures in each cell line; **i**) Tumour mutation burden (TMB) derived from targeted capture sequencing of 57-gene panel and hence abbreviated as 'Surrogate TMB', in 77 paired samples. Briefly, all somatic SNVs or, InDels observed per sample, across the gene-panel are summed to derive surrogate TMB.

### Supplementary_Figure 4. WES and RNA sequencing correlations for *RASSF7*, *RB1* and *SUFU* in primary and replication datasets

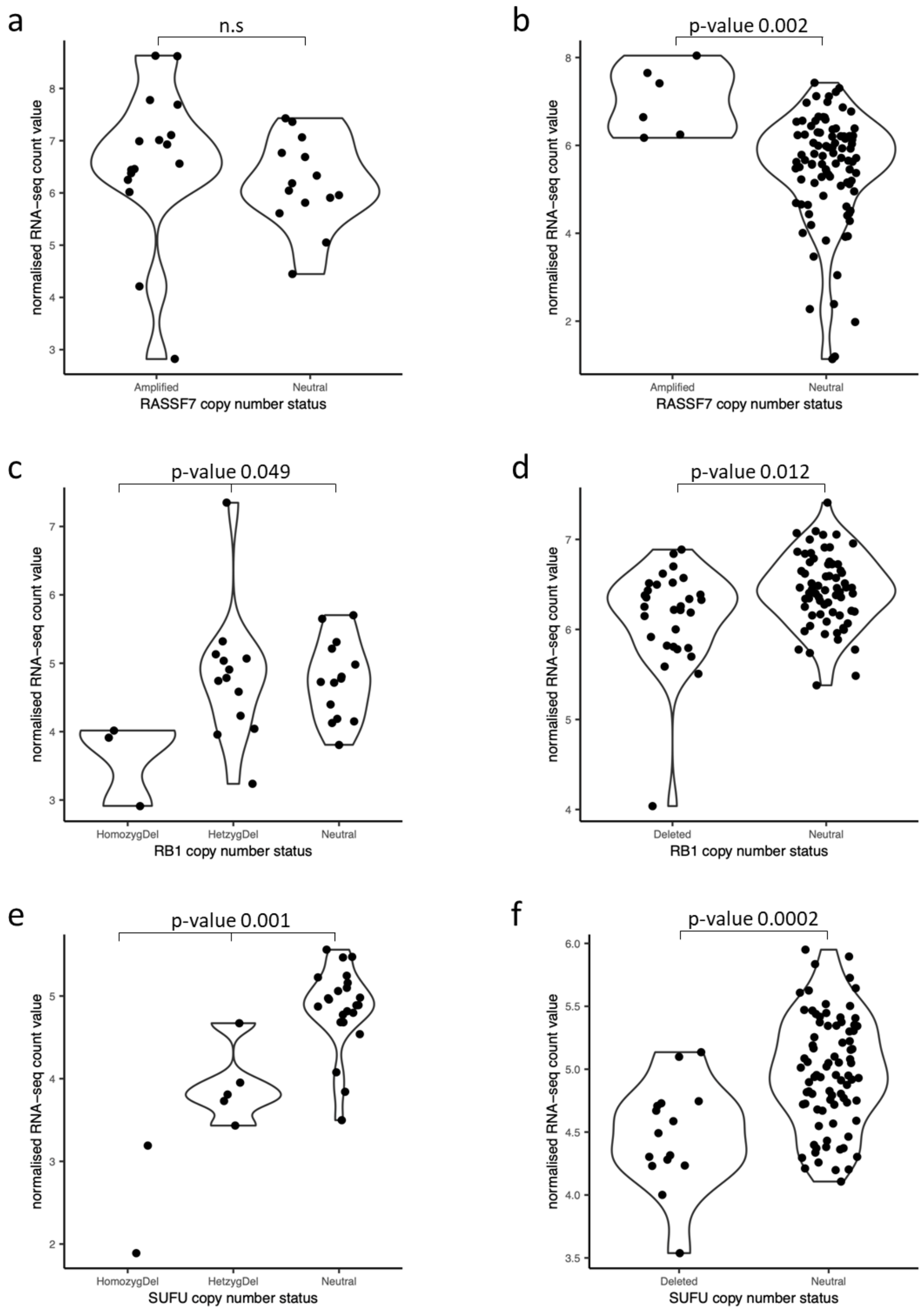

Copy-number profiles from SNP array (NCMR, n=30) (panels **a**, **c** and **e**) or WES (Bueno *et al*., n=98) (**b**, **d** and **f**). were correlated with gene expression from RNA-sequencing. Homozygous vs. heterozygous deletions of *RB1* and *SUFU* loci could be predicted from SNP array data, but we could not discriminate homozygous vs. heterozygous amplification of *RASSF7*. In the Bueno *et al.* WES data only presence of absence of CNAs could be called. Kruskal-Wallis tests were applied to compare expression differences between three classes, and two classes were compared by Mann-Whitney tests. In each case the Bueno *et al*. results confirm the presence of correlations between CNA status and transcript abundance.

### Supplementary_Figure 5. Oncoplot for WGS of mesothelioma primary cells

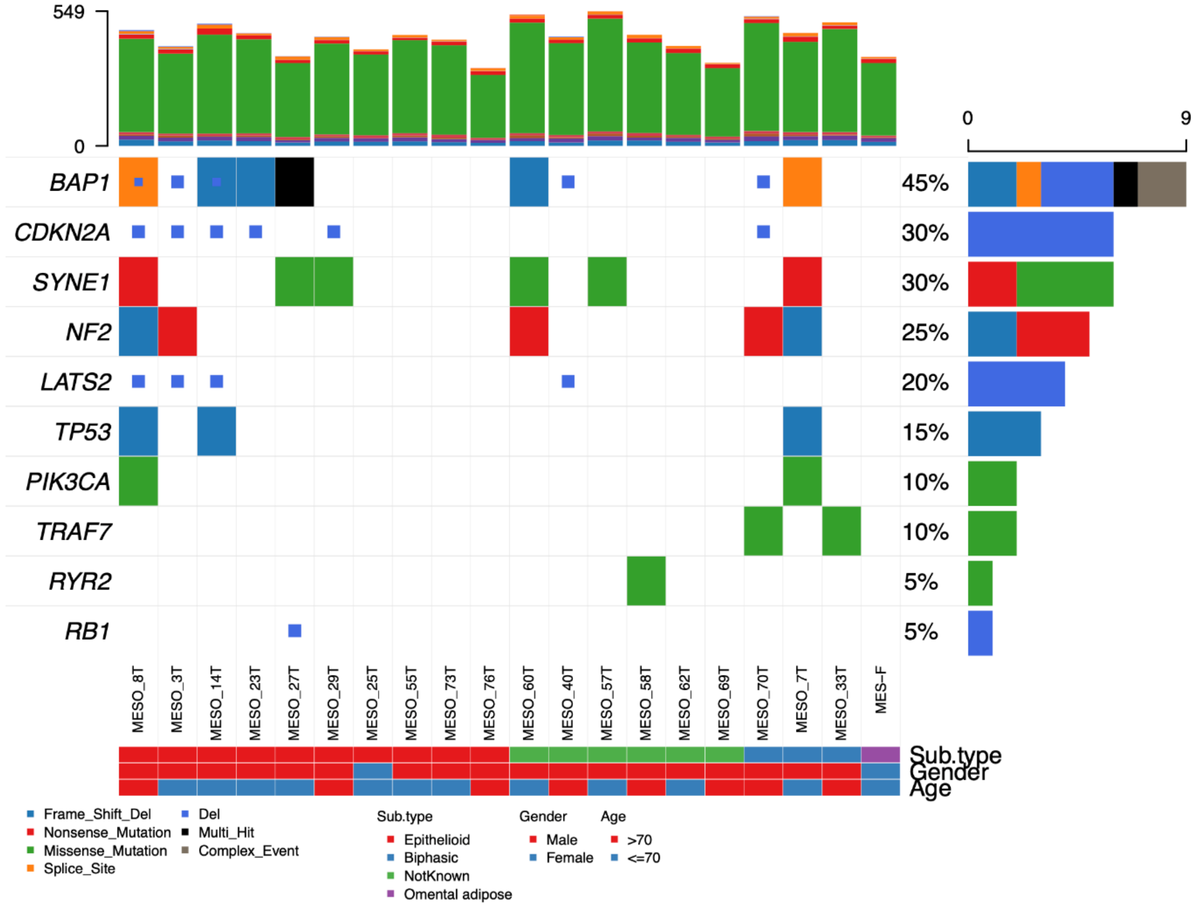

Oncoplot showing alterations in mesothelioma in whole-genome sequenced primary cell lines (19 mesothelioma and one mesothelial primary cell).

### Supplementary_Figure 6. Effect of mutations in DNA damage repair related genes

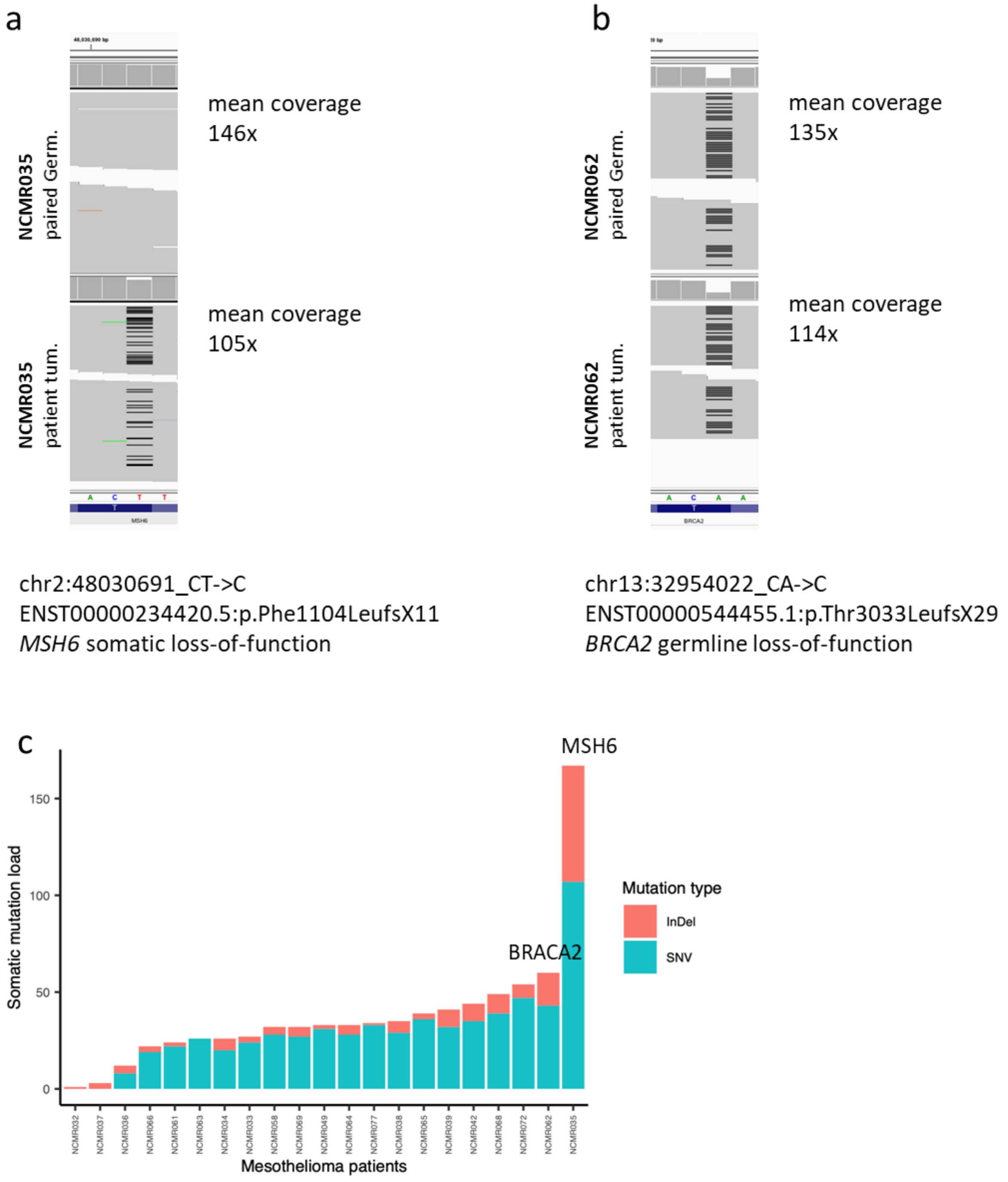

**a** and **b**) IGV (Integrated Genome Viewer) snapshots of matching genomic positions in patient tumour and the paired germline whole-exome sequencing (WES) BAM files. Upper panel shows germline somatic mutational load ((somatic SNV + InDels), lower panel shows tumour mutational load. **a**) is from a patient with somatic MSH6 loss and **b)** is from a patient with germline BRACA2 loss. **c**) Tumour somatic mutational in tumour undergoing WES: BRACA2 and MHS6 mutations are at the extreme of the load spectrum
